# Satisfaction with Life in relation to Sleep Health among a Nationally Representative Sample of U.S. Adults

**DOI:** 10.1101/2025.08.29.25334687

**Authors:** Bethany T. Ogbenna, Symielle A. Gaston, Wensu Zhou, Christopher Payne, W. Braxton Jackson, Chandra L. Jackson

## Abstract

**Purpose:** To investigate associations between life satisfaction and sleep health among adults in the United States.

**Methods:** We analyzed cross-sectional, nationally-representative data from the 2022 National Health Interview Survey. Life satisfaction was dichotomized as ‘very satisfied/satisfied’ vs. ‘dissatisfied/very dissatisfied.’ Sleep duration was defined as ‘recommended’ vs. ‘short’ (≥7 vs.<7 hours), frequent insomnia symptoms as difficulty falling/staying asleep: ‘yes’ [most days/every day to either] vs. ‘no’ [never/some days for both]), and restorative sleep as feeling well rested in the past 30 days: ‘yes’ [never/some days] vs. ‘no’ [most days/every day]. Using survey-weighted Poisson regression with robust variance adjusting for confounders, we estimated prevalence ratios (aPR) and 95% confidence intervals (CI) overall and by age, sex, race, and ethnicity to test for effect modification.

**Results:** Among 25,090 adults (mean age of 48.1±0.17 years; 54% women), 96.0% reported life satisfaction with comparable prevalence across age: 18-30 years [96.3%], 31-49 years [96.6%], and ≥50 years [95.3%]; and among men [95.8%] along with women [96.1%]. Prevalence by race and ethnicity ranged from 93.5% [non-Hispanic (NH)-Multiracial/other] to 98.3% [NH-Asian]. Life satisfaction vs. dissatisfaction was associated with recommended sleep duration (aPR:1.14 [95% CI:1.07-1.21]), restorative sleep (aPR:1.61 [95% CI:1.45 −1.79]), and infrequent insomnia symptoms (aPR:1.25 [95% CI:1.16-1.33]) even after further adjustment. Although life satisfaction varied by age, sex, race, and ethnicity, they did not modify associations between life satisfaction and sleep.

**Conclusions:** Life satisfaction was associated with recommended sleep duration, infrequent insomnia symptoms, and restorative sleep. Pathways underlying the life satisfaction-sleep relationship should be identified to inform interventions.

## Introduction

The National Sleep Foundation (NSF) conducted a ‘Sleep in America®’ poll and found adults in the United States (U.S.) with good vs. poor sleep —defined as meeting NSF sleep duration recommendations, reporting sleep satisfaction, and having no trouble falling asleep—were significantly more likely to report flourishing or an optimal state of well-being [1]. With a critical role in shaping overall well-being, sleep is increasingly recognized as a pillar of mental, emotional, and physical health by, for instance, modulating activity within the hypothalamic-pituitary-adrenal (HPA) axis, reducing cortisol secretion, and promoting synaptic plasticity—mechanisms that collectively support emotional regulation and effective stress management [2–5].

Satisfaction with life – as opposed to flourishing as a dynamic state of realizing potential across life domains – is a different but related indicator of well-being that reflects an individuals’ overall subjective emotional and cognitive assessment of their life along with the degree to which they are content with their life circumstances [6, 7]. Life satisfaction is theorized to include the following domains: a suitable housing and living environment; quality personal relationships and with community members; time to engage in enjoyable and/or meaningful activities (including spiritual); and socioeconomic viability through, for instance, economic stability, job satisfaction, and sufficient work-life integration [8].

Although life satisfaction has been linked to positive health outcomes, its relationship with sleep health remains understudied. The limited published research suggests a bidirectional association: higher life satisfaction may promote favorable sleep through reduced stress, healthier habits (e.g., physical activity; nutrition), and a more positive outlook, while adequate sleep duration, quality, and timing may enhance life satisfaction by supporting emotional regulation, mitigating anxiety and negative mood, and promoting both physical health and cognitive function [9–13]. With few studies conducted in the U.S., mainly international studies have reported associations between higher life satisfaction and longer sleep duration or better sleep quality [11, 14–18]. For example, studies in Germany and China found life satisfaction was associated with fewer sleep complaints, while research in Finland and the Czech Republic emphasized the role of sleep quality [16–19]. However, these studies often focus on middle-aged or older adults, and have limited generalizability to younger, more heterogenous populations like the U.S. [19–21].

Understanding the association between life satisfaction and sleep is important among U.S. adults with varying social characteristics for several reasons. There is known variation in access to health-promoting resources and differential life experiences across sociodemographic groups [22, 23]. For instance, life satisfaction may increase with age but can decline in later life due to chronic conditions or social isolation [24]. While women often report higher life satisfaction than men, women also experience more caregiving roles, depression and insomnia symptoms, and financial stress due to a higher likelihood of having a low-income or living in poverty [25]. Furthermore, racial and ethnic differences are well-documented, with non-Hispanic (NH)-Black and Hispanic adults reporting lower life satisfaction than NH-White adults, potentially due to, on average, lower access to high-quality education and employment opportunities that can lead to lower socioeconomic status and psychological stress [26, 27]. Moreover, short sleep duration is more prevalent among certain racial and ethnic groups, including NH-Black and NH-Pacific Islander, compared to NH-White adults [28, 29]. However, no prior studies, to our knowledge, have assessed the relationship between life satisfaction and sleep health across a large, nationally-representative sample of the U.S. population. Variation by age, sex, race, and ethnicity have also not been studied.

To address these gaps, we determined: (1) the prevalence of life satisfaction overall and by age, sex, and race along with ethnicity; (2) cross-sectional associations between life satisfaction and sleep health overall and within these groups; and (3) differences in sleep health among racial and ethnic groups reporting life satisfaction compared to NH-White adults reporting life satisfaction. We further assessed these associations within age- and sex-specific groups (e.g., comparing Hispanic/Latino adults aged 18–30 years reporting satisfaction or dissatisfaction with NH-White adults reporting satisfaction). We hypothesized that life satisfaction would be more prevalent among younger versus older adults, women versus men, and NH-White versus other racial and ethnic groups. We also hypothesized that life satisfaction would be associated with recommended sleep duration, infrequent insomnia symptoms, and restorative sleep, with stronger associations among younger adults, women, and NH-White adults relative to their counterparts. Finally, we expected Hispanic/Latino, NH-American Indian/Alaska Native, NH-Asian, NH-Black, and NH-multiracial/Other adults reporting life satisfaction to have lower prevalence of recommended sleep, infrequent insomnia symptoms, and restorative sleep compared to NH-White adults with life satisfaction, and even lower prevalence among these groups when reporting life dissatisfaction.

## Methods

### Data source: The National Health Interview Survey

The National Health Interview Survey (NHIS) is a nationally representative household interview survey of non-institutionalized U.S. population. The NHIS uses a complex, multistage probability sample design that incorporates stratification, clustering, and oversampling of certain subgroups (e.g., elderly). The survey is conducted annually via face-to-face interviews with telephone follow-ups and self-administered questionnaires, and collects information on sociodemographic, health behaviors, chronic conditions, and healthcare access and utilization. Additional details on the sampling design and study description were previously described [30, 31]. Informed consent was obtained from all participants by the National Health Interview Survey (NHIS). The Institutional Review Board (IRB) of the National Institute of Environmental Health Sciences (NIEHS) determined that approval was not required for the use of publicly available, de-identified secondary data analysis.

### Study population

Using the Integrated Public Use Microdata Series (IPUMS), we obtained cross-sectional data from the 2022 NHIS survey composed of 35,115 adults [32]. Participants were excluded if missing data for the following: all sleep measures (n=1,044), life satisfaction (n=58), age (n=56), sex (n=3), self-identified race and ethnicity (n=0), or potential confounders (n=1,400) yielding a total of 2,561 excluded from the sample (7.3%). The final analytic sample included 25,090 adults (Supplemental Figure 1). Participants excluded from the analytic sample were more likely to: be women, identify as NH-Black; attain ≤ high school degree; live in the South; and report lifetime abstinence from alcohol, physically inactivity, fair/poor health, and recommended sleep duration. Excluded participants were less likely to be employed, married/living with a partner, and ever have depression (Supplemental Table 1).

### Exposure assessment: Satisfaction with life

Satisfaction with life was measured via self-reported questionnaire by asking participants, “In general, how satisfied are you with your life? Would you say very satisfied, satisfied, dissatisfied, or very dissatisfied?” Responses were dichotomized as ‘very satisfied/satisfied’ vs. ‘dissatisfied/very dissatisfied.

### Outcome assessment: Sleep duration and disturbances

Participants were asked to report their sleep duration by responding to the question, “On average, how many hours of sleep do you get in a 24-hour period?” Responses were recorded as whole numbers, with durations of ≥ 30 minutes rounded up to the nearest hour and durations < 30 minutes rounded down to nearest hour. Responses were categorized as recommended (≥7 hours) vs. < 7 hours sleep duration based on the guidelines from the American Academy of Sleep Medicine and Sleep Research Society [33]. Infrequent insomnia symptoms were measured by participants’ responding to two questions: 1) “During the past 30 days, how often did you have trouble falling asleep?” and 2) “How often did you have trouble staying asleep?” Responses options included “never”, “some days”, “most days” or “every day. Infrequent insomnia symptoms were dichotomized as ‘yes’ if participants responded “never or “some days” to both questions vs. ‘no’ if participants responded “most days” or “every day” to either. Restorative sleep was measured using the question, “During the past 30 days, how often did you wake up feeling well-rested?” with responses dichotomized as ‘yes’ [most days/every day] vs. ‘no’ [never/some days].

### Potential confounders

Potential confounders were identified *a priori* from prior literature and included the following: age (years), sex (man, woman), self-reported race and ethnicity (Hispanic/Latino, NH-American Indian/Alaska Native, NH-Asian, NH-Black/African American, NH-multiracial or other group, NH-White), educational attainment (≤ high school, some college, ≥ college), employment status (employed - currently working, employed - not currently working, not employed - homemaker, not employed - going to school, not employed - retired, not employed - not able to work, not employed - looking for work, not employed - other), and marital status (divorced/widowed, single/no live-in partner, married/living with partner/co-habituating), region of residence (Northeast, Midwest, South, West), cigarette smoking status (never/quit > 12 months prior to interview, former, current), alcohol consumption (current, former, lifetime abstainer), leisure-time physical activity (inactive, insufficiently active, sufficiently active), depression (yes, no), and BMI (underweight [<18.5 kg/m], recommended [18.5-24.9 kg/m], overweight [25-29.9 kg/m], obesity [≥30 kg/m].

### Potential effect measure modifiers

We investigated the following potential effect modifiers: age category (18–30, 31–49, ≥50 years), sex, as well as combined race and ethnicity based on prior literature suggesting these sociodemographic factors may modify satisfaction with life [24, 34, 35].

### Statistical analysis

Descriptive statistics were estimated overall and by life satisfaction (satisfied vs. dissatisfied). We reported sample weighted means (+/- standard errors) for age and age-standardized (consistent with the 2010 U.S. Census population) sample weighted proportions for categorical variables. The NHIS sampling weights used in these analyses account for the inverse probability of selection to reflect the complex survey design and correct for non-response. Poisson regression with robust standard errors [36, 37] was used to estimate adjusted prevalence ratios and 95% confidence intervals (aPR [95%CI]) for associations between life satisfaction and sleep health adjusting for potential confounders in the overall population and stratified by potential modifiers (i.e., age, sex, race and ethnicity), separately. Model 1 was adjusted for age sex, race and ethnicity (when models were not stratified by these variables), marital status, educational attainment, employment status, and general health status. Model 2 was further adjusted for potential mediators: BMI, leisure-time physical activity, smoking status, alcohol consumption, and depression. To test for effect modification, cross-product interaction terms (e.g., life satisfaction*age, life satisfaction*sex, and life satisfaction*race/ethnicity) were included in the overall model containing Model 2 covariates. All racial and ethnic groups were included in analyses, however, due to heterogeneity, impactful results are interpreted in the text based on either magnitude of point estimates or significant alpha testing. All analyses were performed using estimation and post-estimation commands for survey data in Stata, Version 15.1 (Statacorp, College Station, Texas), and a two-sided p-value of 0.05 was used to determine statistical significance. All results in text unless otherwise stated are reported from fully adjusted models (Model 2).

## Results

### Study population Characteristics

Among participants (N = 25,090; 54.0% women), the mean age (SE) was 48 (0.2) years (Table 1). Overall, satisfaction with life was prevalent (95.6%) and comparable between men (95.6%) and women (95.6%) (Table 1) as well as across age groups (Supplemental Table 2) as well as. Satisfaction with life varied by race and ethnicity among Hispanic/Latine (96.5%), NH-American Indian/Alaska Native (93.0%), NH-Asian (98.0%), NH-Black (95.3%), NH-multiracial/Other (93.9%), and NH-White (95.4%) (see Supplemental Table 3). Participants satisfied with life were more likely to complete ≥ college (33.8% vs. 19.2% dissatisfied), be employed (64.6% vs. 41.0% dissatisfied), married (60.2 % vs. 34.6% dissatisfied), live in the southern U.S. region (38.0% vs. 35.4% dissatisfied), never smoke (87.8% vs. 73.9% dissatisfied), consume alcohol (70.2% vs. 64.4% dissatisfied), be sufficiently physically active (48.5% vs. 32.3% dissatisfied), be overweight (34.0% vs. 27.0% dissatisfied), and report good/very good/excellent self-rated general health (87.7% vs. 48.2% dissatisfied). Adults who were satisfied with life had a higher prevalence of favorable sleep outcomes compared to adults who were dissatisfied: 70.3% vs. 54.6% for recommended sleep duration, 77.8% vs. 49.6% for infrequent insomnia symptoms, and 58.3% vs. 25.7% for restorative sleep.

**Table 1.** Study population characteristics, overall and by sex, National Health Interview Survey, 2022, (N=25,090)

|  | Total<br>n=25,090 (100%) |  |  | Men<br>n=11,532 (46.0%) |  |  | Women<br>n=13,558 (54.0%) |  |  |
| --- | --- | --- | --- | --- | --- | --- | --- | --- | --- |
|  | Life satisfaction<sup>a</sup> |  |  | Life satisfaction<sup>a</sup> |  |  | Life satisfaction<sup>a</sup> |  |  |
|  | All<br>n=25,090<br>(100%) | Yes<br>n=23,997<br>(95.6%) | No<br>n=1,093<br>(4.4%) | All<br>n=11,532<br>(100%) | Yes<br>n=11,030<br>(95.6%) | No<br>n=502<br>(4.4%) | All<br>n=13,558<br>(100%) | Yes<br>n=12,967<br>(95.6%) | No<br>n=591<br>(4.4%) |
| Sociodemographic Characteristics |  |  |  |  |  |  |  |  |  |
| Age (years), mean (SE) | 48.1 (.17) | 48.0 (.17) | 50.5 (.75) | 47.4 (.21) | 47.4 (.22) | 48.2 (1.1) | 48.7 (.21) | 48.5 (.21) | 52.8 (.9 |
| 18-30 years | 21.8 | 21.9 | 20.0 | 22.5 | 22.4 | 25.0 | 21.1 | 21.4 | 14.9 |
| 31-49 years | 31.5 | 31.7 | 26.1 | 31.9 | 32.2 | 25.7 | 31.0 | 31.2 | 26.4 |
| ≥ 50 years | 46.7 | 46.4 | 53.9 | 45.6 | 45.4 | 49.3 | 47.9 | 47.4 | 58.7 |
| Race and ethnicity |  |  |  |  |  |  |  |  |  |
| Hispanic/Latine | 16.9 | 17.1 | 13.6 | 16.9 | 17.0 | 13.5 | 17.0 | 17.2 | 13.6 |
| NH-American Indian/Alaska Native | 0.8 | 0.8 | 1.0 | 0.6 | 0.6 | 1.4 | 0.9 | 1.0 | 0.5 |
| NH-Asian | 6.1 | 6.3 | 2.7 | 5.7 | 5.8 | 2.5 | 6.5 | 6.7 | 2.7 |
| NH-Black/African American | 11.2 | 11.1 | 13.5 | 10.1 | 10.0 | 13.4 | 12.2 | 12.2 | 13.5 |
| NH-multiracial or other group<sup>b</sup> | 2.0 | 2.0 | 3.4 | 2.1 | 2.1 | 3.7 | 1.9 | 1.9 | 3.3 |
| NH-White | 62.9 | 62.8 | 65.9 | 64.6 | 64.5 | 65.6 | 61.3 | 61.1 | 66.5 |
| Educational Attainment |  |  |  |  |  |  |  |  |  |
| ≤ High School | 37.2 | 36.8 | 47.3 | 39.1 | 38.7 | 50.5 | 35.1 | 34.8 | 42.9 |
| Some college | 29.6 | 29.5 | 33.6 | 28.3 | 28.1 | 31.9 | 30.9 | 30.8 | 36.1 |
| ≥ College | 33.2 | 33.8 | 19.2 | 32.6 | 33.2 | 17.7 | 33.9 | 34.4 | 21.0 |
| Employment status |  |  |  |  |  |  |  |  |  |
| Employed, currently working | 63.7 | 64.6 | 41.0 | 68.6 | 69.7 | 43.1 | 58.9 | 59.7 | 38.9 |
| Employed, not currently working | 0.6 | 0.6 | 0.9 | 0.6 | 0.6 | 0.7 | 0.6 | 0.6 | 1.0 |
| Not employed, Homemaker | 4.7 | 4.7 | 4.7 | 0.9 | 0.8 | 1.8 | 8.6 | 8.6 | 8.4 |
| Not employed, Going to school | 2.3 | 2.4 | 1.6 | 2.2 | 2.2 | 2.5 | 2.5 | 2.6 | 0.4 |
| Not employed, Retired | 18.9 | 19.0 | 16.8 | 18.0 | 18.1 | 16.0 | 19.7 | 19.8 | 17.5 |
| Not employed, not able to work | 6.0 | 5.1 | 25.0 | 5.7 | 4.9 | 23.8 | 6.2 | 5.3 | 26.0 |
| Not employed, looking for work | 2.2 | 2.0 | 6.3 | 2.5 | 2.3 | 8.0 | 1.9 | 1.8 | 4.5 |
| Not employed, Other | 1.6 | 1.5 | 3.8 | 1.5 | 1.4 | 4.1 | 1.6 | 1.6 | 3.3 |
| Marital status |  |  |  |  |  |  |  |  |  |
| Divorced/widowed | 17.2 | 16.7 | 28.0 | 13.2 | 12.8 | 22.4 | 20.8 | 20.2 | 33.4 |
| Single/no live-in partner | 23.6 | 23.1 | 37.4 | 25.3 | 24.5 | 42.7 | 22.0 | 21.7 | 30.5 |
| Married/living with partner/cohabiting | 59.2 | 60.2 | 34.6 | 61.5 | 62.7 | 34.9 | 57.2 | 58.1 | 36.1 |
| Region of residence |  |  |  |  |  |  |  |  |  |
| Northeast | 17.2 | 17.2 | 17.8 | 17.3 | 17.3 | 17.6 | 17.1 | 17.1 | 17.7 |
| Midwest | 21.0 | 21.0 | 22.1 | 21.4 | 21.4 | 21.2 | 20.7 | 20.6 | 22.7 |
| South | 37.9 | 38.0 | 35.4 | 37.3 | 37.4 | 35.1 | 38.6 | 38.7 | 36.4 |
| West | 23.8 | 23.8 | 24.7 | 24.0 | 24.0 | 26.1 | 23.7 | 23.7 | 23.1 |
| Health Behaviors |  |  |  |  |  |  |  |  |  |
| Smoking status |  |  |  |  |  |  |  |  |  |
| Never/quit>12 months prior | 87.3 | 87.8 | 73.9 | 85.5 | 86.1 | 73.2 | 89.0 | 89.5 | 74.6 |
| Former/quit≤12 months ago | 1.2 | 1.2 | 2.4 | 1.4 | 1.4 | 3.2 | 0.9 | 0.9 | 1.5 |
| Current | 11.5 | 11.1 | 23.7 | 13.0 | 12.6 | 23.5 | 10.1 | 9.6 | 23.9 |
| Alcohol consumption |  |  |  |  |  |  |  |  |  |
| Current (≥1 drink past year) | 69.9 | 70.2 | 64.4 | 72.8 | 73.0 | 67.6 | 67.3 | 67.6 | 62.4 |
| Former (no drinks past year) | 17.0 | 16.7 | 23.8 | 17.2 | 17.0 | 22.5 | 16.8 | 16.5 | 24.4 |
| Lifetime abstinence (<12 drinks in life) | 13.0 | 13.1 | 11.7 | 10.0 | 10.0 | 9.9 | 15.9 | 16.0 | 13.2 |
| Leisure-time physical activity<sup>c</sup> |  |  |  |  |  |  |  |  |  |
| Inactive | 26.5 | 25.8 | 44.3 | 24.7 | 23.9 | 42.6 | 28.3 | 27.5 | 46.5 |
| Insufficiently active | 25.6 | 25.7 | 23.4 | 22.7 | 22.8 | 20.5 | 28.5 | 28.6 | 26.4 |
| Sufficiently active | 47.8 | 48.5 | 32.3 | 52.6 | 53.3 | 36.9 | 43.2 | 43.9 | 27.0 |
| Usual sleep duration |  |  |  |  |  |  |  |  |  |
| Short (<7 hours) | 30.3 | 29.7 | 45.4 | 29.9 | 29.3 | 44.0 | 30.6 | 30.0 | 46.9 |
| Recommended (≥7 hours) | 69.7 | 70.3 | 54.6 | 70.1 | 70.7 | 56.0 | 69.4 | 70.0 | 53.1 |
| Infrequent Insomnia symptoms (yes)<sup>d</sup> | 76.7 | 77.8 | 49.6 | 80.1 | 81.2 | 53.0 | 73.4 | 74.5 | 46.1 |
| Restorative sleep (yes)<sup>e</sup> | 57.0 | 58.3 | 25.7 | 60.6 | 62.1 | 27.0 | 53.3 | 54.5 | 24.3 |
| Clinical Characteristics |  |  |  |  |  |  |  |  |  |
| Ever had depression (yes)<sup>f</sup> | 18.5 | 16.9 | 55.2 | 13.2 | 11.6 | 49.0 | 23.6 | 22.0 | 62.2 |
| Body mass index category |  |  |  |  |  |  |  |  |  |
| Underweight (<18.5 kg/m<sup>2</sup>) | 1.7 | 1.6 | 2.8 | 1.1 | 1.1 | 1.3 | 2.2 | 2.1 | 4.3 |
| Recommended (18.5-<25 kg/m<sup>2</sup>) | 31.2 | 31.3 | 28.2 | 27.0 | 26.9 | 29.2 | 35.3 | 35.6 | 26.7 |
| Overweight (25-<30 kg/m<sup>2</sup>) | 33.7 | 34.0 | 27.0 | 38.8 | 39.0 | 34.2 | 28.8 | 29.2 | 19.4 |
| Obesity (≥30 kg/m<sup>2</sup>) | 33.4 | 33.0 | 42.0 | 33.1 | 33.0 | 35.4 | 33.7 | 33.1 | 49.6 |
| General health status |  |  |  |  |  |  |  |  |  |
| Fair/poor | 13.9 | 12.3 | 51.8 | 13.6 | 12.0 | 50.4 | 14.3 | 12.6 | 53.8 |
| Good/very good/excellent | 86.1 | 87.7 | 48.2 | 86.4 | 88.0 | 49.6 | 85.7 | 87.4 | 46.2 |

Abbreviations: SE (standard error), NH (non-Hispanic)
Note: Data are presented as column percentages or means and standard errors. Percentages may not sum to 100 due to missing rounding. All estimates are weighted for the survey's complex sampling design. All estimates are age-standardized to the U.S. 2020 population, except for age.
<sup>a</sup> Life satisfaction was ascertained based on the questions, 'In general, how satisfied are you with your life? Are you very satisfied, satisfied, dissatisfied, or very dissatisfied?'. Life satisfaction was dichotomized as yes (a response of 'satisfied' or 'very satisfied') vs. no (a response of 'very dissatisfied' or 'dissatisfied').
<sup>b</sup> NH-Other single and multiple races and 'Other group' is defined as persons identifying with racial groups not explicitly listed in the standard categories.
<sup>c</sup> Leisure-time physical activity was defined using the 2018 Health and Human Services Physical Activity guidelines which state "recommend that adults complete at least 150 minutes to 300 minutes of moderate-intensity activity, or 75 minutes to 150 minutes of vigorous-intensity aerobic activity per week, as well as moderate or greater intensity muscle strengthening activities on two or more days a week."
<sup>d</sup> Infrequent insomnia symptoms (yes) defined as difficulty falling or staying asleep most days/every day.
<sup>e</sup> Restorative sleep (yes) defined as never/some days waking up feeling rested in the past 30 days.
<sup>f</sup> Depression was defined by the 2019 Field Representative's Manual as a major depressive disorder or as clinical depression that is a common but serious mood disorder. It causes severe symptoms that affect how you feel, think, and handle daily activities, such as sleeping, eating, or working."

### Life satisfaction and sleep health in the overall population

Overall, life satisfaction versus dissatisfaction was associated with a 14% higher prevalence of recommended sleep duration (aPR: 1.14, 95% CI: 1.07, 1.21), a 25% higher prevalence of infrequent insomnia symptoms (aPR:1.25 95% CI:1.16, 1.33), and a 61% higher prevalence of restorative sleep (aPR:1.61, 95% CI:1.45, 1.79) (Table 2).

**Table 2.**
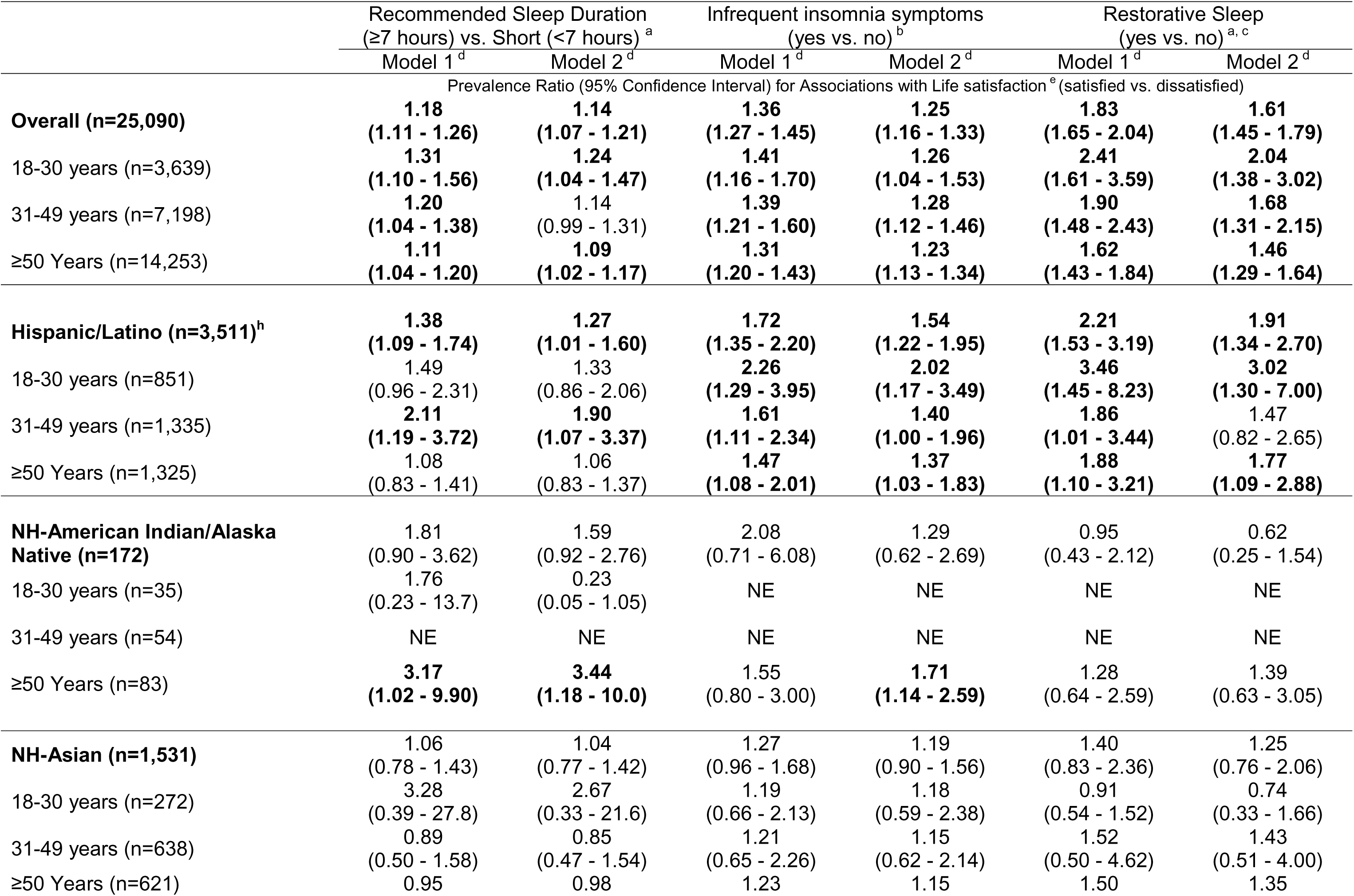
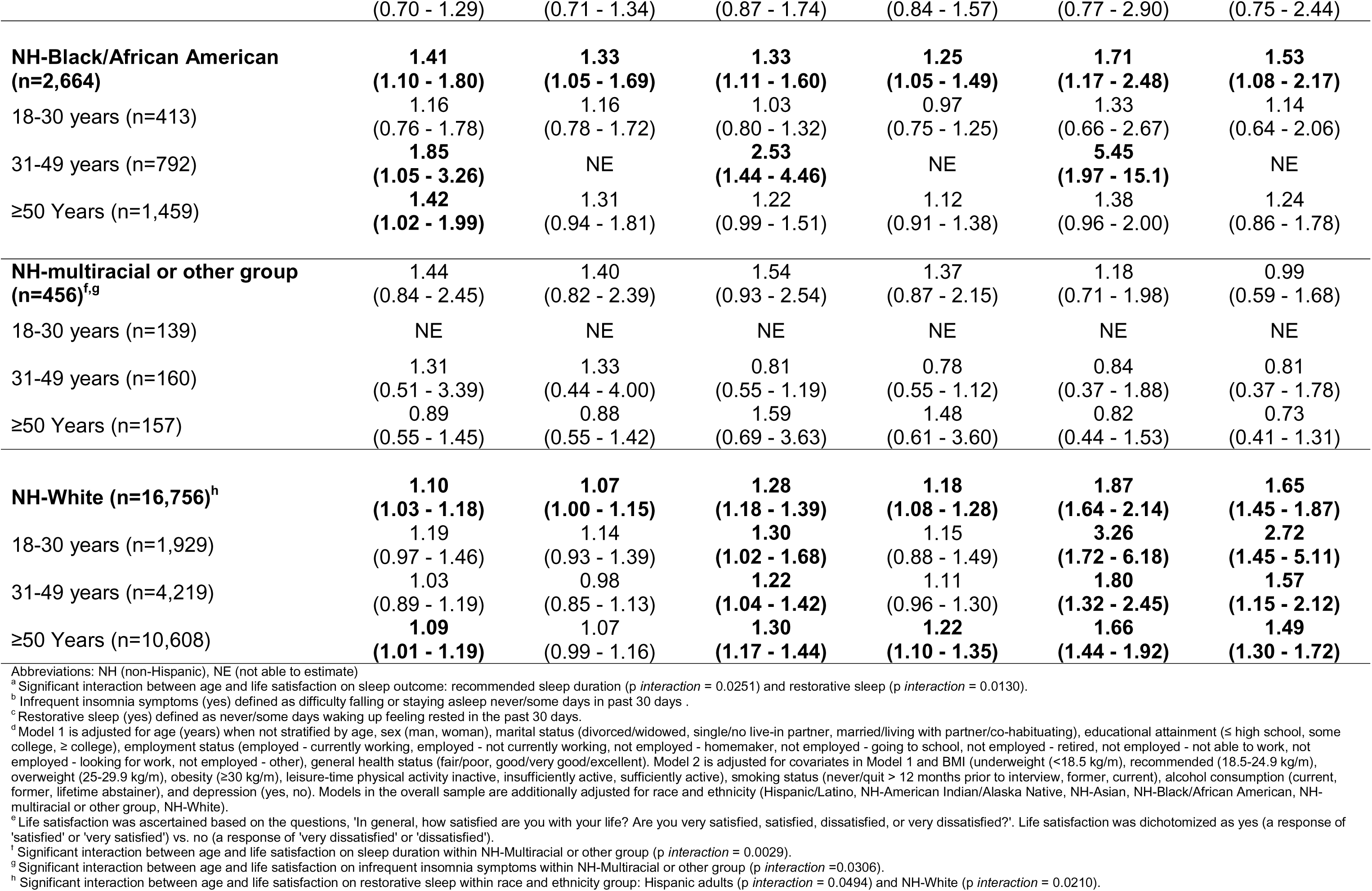
Prevalence ratios for associations between life satisfaction and sleep, overall and stratified by race along with ethnicity and age, National Health Interview Survey, 2022, (N=25,090)

### Life satisfaction and sleep health by age

Potential evidence of effect modification by age was observed for the associations between life satisfaction and both recommended sleep duration and restorative sleep based on p-values for cross-product interaction terms, however; confidence intervals for age-specific estimates largely overlapped. Life satisfaction and age on recommended sleep duration was observed, with stronger associations among adults 18-30 years (aPR_18-30 years_:1.24, 95% CI:1.04,1.47), followed by adults 31-49 years (aPR_31-49 years_:1.14, 95% CI:0.99,1.31), and adults ≥50 years (aPR _≥ 50 years_:1.09, 95% CI:1.02, 1.17; *p* _life satisfaction*age_ = 0.025) (see Table 2). Life satisfaction and age on restorative sleep was observed among adults 18-30 years (aPR_18-30 years_ :2.04, 95% CI:1.38, 3.02) followed by adults 31-49 years (aPR_31-49 years_ :1.68, 95% CI:1.31,2.15), and adults≥ 50 years (aPR _≥ 50 years_ :1.46, 95% CI:1.29, 1.64; *p* _life satisfaction*age_ = 0.013).

### Life satisfaction and sleep health by sex and race and ethnicity

Neither sex (Table 3) nor race and ethnicity (Table 2) modified the associations between life satisfaction and sleep health dimensions among the overall population (*p* _life satisfaction*race and ethnicity_ > 0.05).

**Table 3.** Prevalence ratios for associations between life satisfaction and sleep, overall and stratified by race along with ethnicity and sex, National Health Interview Survey, 2022, (N=25,090)

|  | Recommended Sleep Duration (≥7 hours) vs. Short (<7 hours) |  | Infrequent insomnia symptoms (yes vs. no)<sup>a</sup> |  | Restorative Sleep (yes vs. no)<sup>b</sup> |  |
| --- | --- | --- | --- | --- | --- | --- |
|  | Model 1<sup>c</sup> | Model 2<sup>c</sup> | Model 1<sup>c</sup> | Model 2<sup>c</sup> | Model 1<sup>c</sup> | Model 2<sup>c</sup> |
|  | Prevalence Ratio (95% Confidence Interval) for Associations with Life satisfaction<sup>d</sup> (satisfied vs. dissatisfied) |  |  |  |  |  |
| **Overall (n=25,090)** | **1.18**<br>**(1.11 - 1.26)** | **1.14**<br>**(1.07 - 1.21)** | **1.36**<br>**(1.27 - 1.45)** | **1.25**<br>**(1.16 - 1.33)** | **1.83**<br>**(1.65 - 2.04)** | **1.61**<br>**(1.45 - 1.79)** |
| Men (n=11,532) | 1.19<br>**(1.09 - 1.31)** | 1.14<br>**(1.04 - 1.25)** | 1.35<br>**(1.22 - 1.50)** | 1.25<br>**(1.13 - 1.38)** | 1.95<br>**(1.66 - 2.29)** | 1.71<br>**(1.46 - 2.00)** |
| Women (n=13,558) | 1.17<br>**(1.07 - 1.28)** | 1.13<br>**(1.03 - 1.24)** | 1.36<br>**(1.22 - 1.51)** | 1.24<br>**(1.12 - 1.38)** | 1.72<br>**(1.48 - 2.00)** | 1.52<br>**(1.31 - 1.76)** |
| **Hispanic/Latino (n=3,511)** | **1.38**<br>**(1.09 - 1.74)** | **1.27**<br>**(1.01 - 1.60)** | **1.72**<br>**(1.35 - 2.20)** | **1.54**<br>**(1.22 - 1.95)** | **2.21**<br>**(1.53 - 3.19)** | **1.91**<br>**(1.34 - 2.70)** |
| Men (n=1,610) | 1.49<br>**(1.07 - 2.08)** | 1.39<br>**(1.00 - 1.94)** | 1.78<br>**(1.25 - 2.53)** | 1.60<br>**(1.13 - 2.27)** | 2.32<br>**(1.32 - 4.07)** | 2.01<br>**(1.16 - 3.48)** |
| Women (n=1,901) | 1.23<br>**(0.92 - 1.66)** | 1.12<br>**(0.84 - 1.51)** | 1.62<br>**(1.17 - 2.24)** | 1.45<br>**(1.07 - 1.97)** | 2.07<br>**(1.25 - 3.43)** | 1.75<br>**(1.08 - 2.82)** |
| **NH-American Indian/Alaska Native (n=172)** | 1.81<br>**(0.90 - 3.62)** | 1.59<br>**(0.92 - 2.76)** | 2.08<br>**(0.71 - 6.08)** | 1.29<br>**(0.62 - 2.69)** | 0.95<br>**(0.43 - 2.12)** | 0.62<br>**(0.25 - 1.54)** |
| Men (n=71) | 1.22<br>**(0.62 - 2.42)** | 0.80<br>**(0.25 - 2.51)** | 7.20<br>**(0.67 - 77.8)** | 3.34<br>**(0.57 - 19.7)** | 1.57<br>**(0.57 - 4.27)** | 1.53<br>**(0.44 - 5.32)** |
| Women (n=101) | 1.74<br>**(1.09 - 2.79)** | NE | 0.67<br>**(0.43 - 1.05)** | 0.68<br>**(0.36 - 1.30)** | 0.74<br>**(0.37 - 1.48)** | NE |
| **NH-Asian (n=1,531)** | 1.06<br>**(0.78 - 1.43)** | 1.04<br>**(0.77 - 1.42)** | 1.27<br>**(0.96 - 1.68)** | 1.19<br>**(0.90 - 1.56)** | 1.40<br>**(0.83 - 2.36)** | 1.25<br>**(0.76 - 2.06)** |
| Men (n=669) | 1.12<br>**(0.75 - 1.68)** | 1.14<br>**(0.77 - 1.69)** | 1.29<br>**(0.87 - 1.90)** | 1.17<br>**(0.76 - 1.79)** | 1.54<br>**(0.69 - 3.44)** | 1.31<br>**(0.58 - 2.92)** |
| Women (n=862) | 0.91<br>**(0.60 - 1.37)** | 0.89<br>**(0.59 - 1.34)** | 1.21<br>**(0.82 - 1.78)** | 1.17<br>**(0.83 - 1.64)** | 1.26<br>**(0.62 - 2.58)** | 1.16<br>**(0.59 - 2.29)** |
| **NH-Black/African American (n=2,664)** | **1.41**<br>**(1.10 - 1.80)** | **1.33**<br>**(1.05 - 1.69)** | **1.33**<br>**(1.11 - 1.60)** | **1.25**<br>**(1.05 - 1.49)** | **1.71**<br>**(1.17 - 2.48)** | **1.53**<br>**(1.08 - 2.17)** |
| Men (n=1,083) | 1.56<br>**(1.09 - 2.24)** | 1.45<br>**(1.03 - 2.04)** | 1.32<br>**(1.04 - 1.67)** | 1.26<br>**(1.01 - 1.59)** | 1.94<br>**(1.10 - 3.43)** | 1.77<br>**(1.04 - 3.00)** |
| Women (n=1,581) | 1.26<br>**(0.89 - 1.77)** | 1.20<br>**(0.86 - 1.69)** | 1.35<br>**(1.02 - 1.79)** | 1.24<br>**(0.94 - 1.64)** | 1.48<br>**(0.89 - 2.46)** | 1.28<br>**(0.80 - 2.05)** |
| **NH-multiracial or other group**<br>**(n=456)** | **1.44**<br>(0.84 - 2.45) | **1.40**<br>(0.82 - 2.39) | **1.54**<br>(0.93 - 2.54) | **1.37**<br>(0.87 - 2.15) | **1.18**<br>(0.71 - 1.98) | **0.99**<br>(0.59 - 1.68) |
| Men (n=226) | 0.92<br>(0.56 - 1.50) | 0.86<br>(0.54 - 1.38) | 1.57<br>(0.75 - 3.29) | 1.42<br>(0.75 - 2.72) | 1.14<br>(0.59 - 2.20) | 0.98<br>(0.56 - 1.74) |
| Women (n=230) | **2.92**<br>**(1.05 - 8.12)** | **3.13**<br>**(1.17 - 8.39)** | 1.52<br>(0.79 - 2.94) | 1.28<br>(0.73 - 2.22) | 1.39<br>(0.62 - 3.14) | 1.14<br>(0.45 - 2.89) |
| **NH-White (n=16,756)** | **1.10**<br>**(1.03 - 1.18)** | **1.07**<br>**(1.00 - 1.15)** | **1.28**<br>**(1.18 - 1.39)** | **1.18**<br>**(1.08 - 1.28)** | **1.87**<br>**(1.64 - 2.14)** | **1.65**<br>**(1.45 - 1.87)** |
| Men (n=7,873) | 1.10<br>(0.99 - 1.21) | 1.06<br>(0.96 - 1.16) | **1.25**<br>**(1.12 - 1.40)** | **1.16**<br>**(1.04 - 1.30)** | **1.98**<br>**(1.64 - 2.41)** | **1.73**<br>**(1.44 - 2.09)** |
| Women (n=8,883) | **1.12**<br>**(1.01 - 1.23)** | 1.09<br>(0.98 - 1.20) | **1.31**<br>**(1.15 - 1.49)** | **1.20**<br>**(1.05 - 1.37)** | **1.78**<br>**(1.47 - 2.14)** | **1.57**<br>**(1.30 - 1.90)** |

Abbreviations: NH (non-Hispanic), NE (not able to estimate)
<sup>a</sup> Infrequent insomnia symptoms (yes) defined as difficulty falling or staying asleep never/some days in past 30 days
<sup>b</sup> Restorative sleep (yes) defined as never/some days waking up feeling rested in the past 30 days.
<sup>c</sup> Model 1 is adjusted for age (years) when not stratified by age, sex (man, woman), marital status (divorced/widowed, single/no live-in partner, married/living with partner/co-habituating), educational attainment ( ≤ high school, some college, ≥ college), employment status (employed - currently working, employed - not currently working, not employed - homemaker, not employed - going to school, not employed - retired, not employed - not able to work, not employed - looking for work, not employed - other), general health status (fair/poor, good/very good/excellent). Model 2 is adjusted for covariates in Model 1 and body mass index (underweight ( $<18.5$ kg/m), recommended (18.5-24.9 kg/m), overweight (25-29.9 kg/m), obesity ( $\geq 30$ kg/m), leisure-time physical activity inactive, insufficiently active, sufficiently active), smoking status (never/quit $> 12$ months prior to interview, former, current), alcohol consumption (current, former, lifetime abstainer), and depression (yes, no). Models in the overall sample are additionally adjusted for race and ethnicity (Hispanic/Latino, NH-American Indian/Alaska Native, NH-Asian, NH-Black/African American, NH-Multiracial or other group, NH-White)
<sup>d</sup> Life satisfaction was ascertained based on the questions, 'In general, how satisfied are you with your life? Are you very satisfied, satisfied, dissatisfied, or very dissatisfied?'. Life satisfaction was dichotomized as yes (a response of 'satisfied' or 'very satisfied') vs. no (a response of 'very dissatisfied' or 'dissatisfied').

### Various interrelated life satisfaction-sleep associations with age, sex, and race along with ethnicity

#### Life satisfaction and sleep health by age as well as race and ethnicity

Despite overlapping confidence intervals across age groups, the associations between life satisfaction and restorative sleep among NH-White adults appeared to attenuate as age increased (aPR_18-30 years_= 2.72, 95% CI:1.45,5.11; aPR_31-49 years_=1.57, 95% CI:1.15,2.12; aPR _≥ 50 years_=1.49, 95% CI:1.30, 1.72; *p* _life satisfaction*age within NH-White adults_ = 0.021) (Table 2). Age within other racial and ethnic groups did not modify associations with life satisfaction on recommended sleep duration and infrequent insomnia symptoms. (Table 2).

#### Life satisfaction and sleep health by sex as well as race and ethnicity

Within racial and ethnic groups, sex did not modify associations between life satisfaction in relation to recommended sleep duration, infrequent insomnia symptoms, and restorative sleep (Table 3).

#### Life satisfaction among various racial and ethnic groups compared to NH-White adults satisfied with life, overall

Compared to NH-White adults with life satisfaction, Hispanic/Latino adults with life satisfaction had a similar prevalence of recommended sleep duration (aPR=0.98, 95% CI: 0.95–1.01) and restorative sleep (aPR=0.98, 95% CI: 0.95–1.02), as well as an 8% higher prevalence of infrequent insomnia symptoms (aPR=1.08, 95% CI: 1.06–1.11) (Table 4). In contrast, Hispanic/Latino adults reporting life dissatisfaction had significantly lower prevalence of all three sleep outcomes: 27% lower for recommended sleep duration (aPR=0.73, 95% CI: 0.59–0.91), 31% lower for absence of frequent insomnia symptoms (aPR=0.69, 95% CI: 0.55–0.88), and 49% lower for restorative sleep (aPR=0.51, 95% CI: 0.36–0.72). Compared to NH-White adults who were satisfied with life, NH-Black adults with life satisfaction had a 14% lower prevalence of recommended sleep duration (aPR=0.86, 95% CI: 0.83– 0.90) and slightly lower prevalence of restorative sleep (aPR=0.96, 95% CI: 0.92–1.00). NH-Black adults reporting life dissatisfaction had a 36% lower prevalence of recommended sleep duration (aPR=0.64, 95% CI: 0.50–0.82) and a 37% lower prevalence of restorative sleep (aPR=0.63, 95% CI: 0.45–0.88).

**Table 4.**
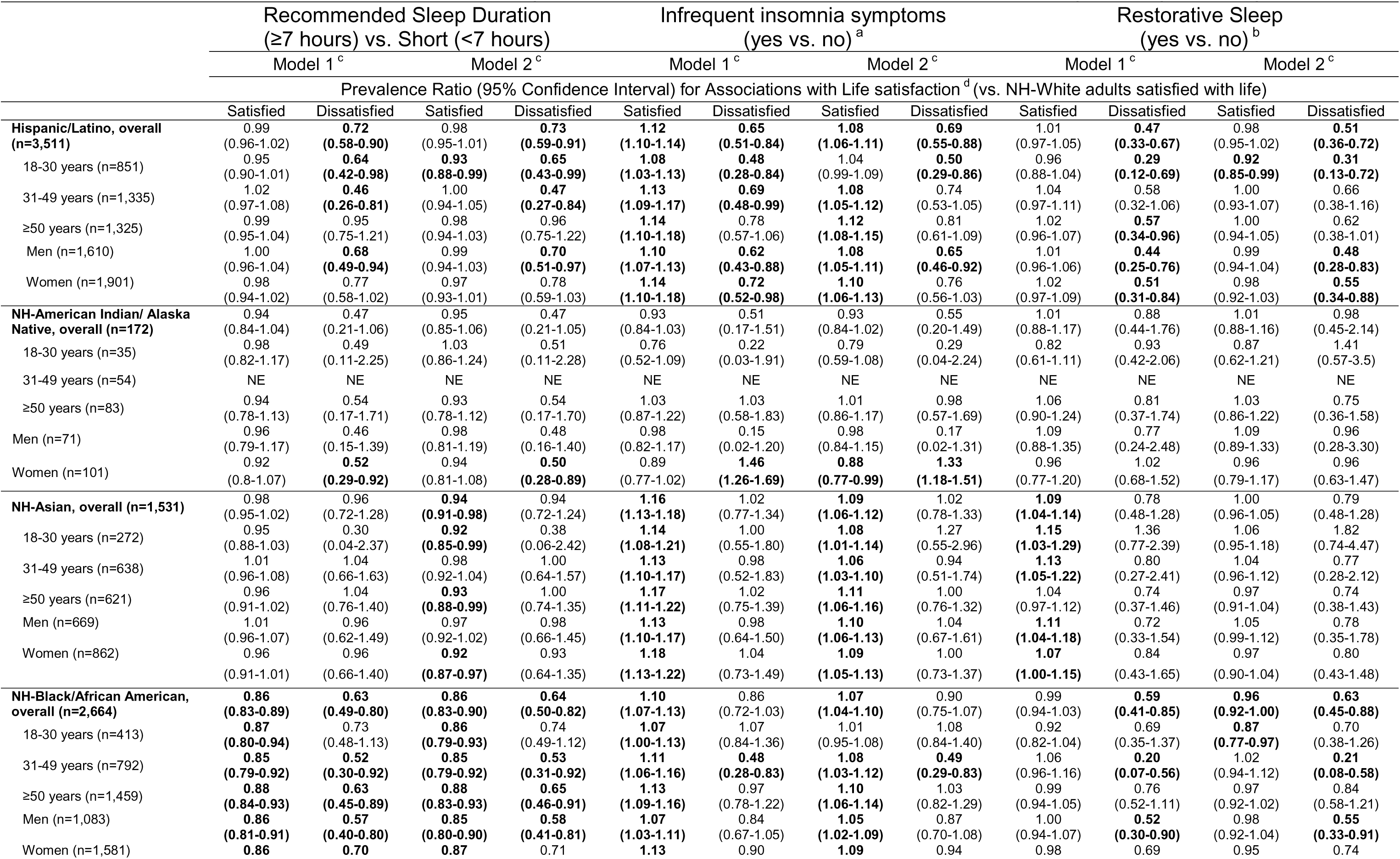
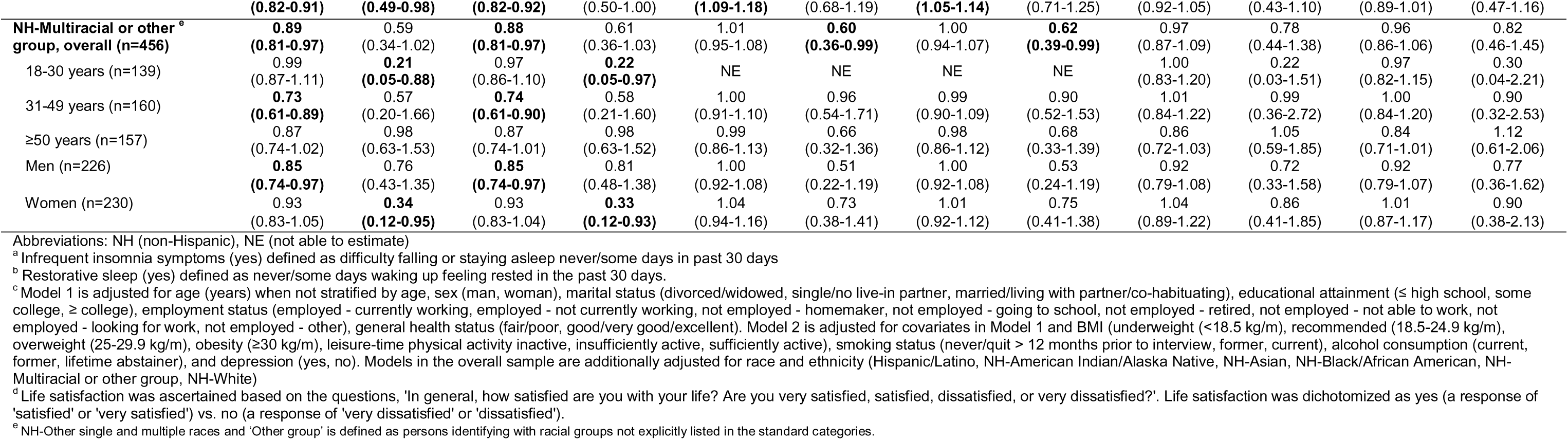
Prevalence ratios of sleep duration and sleep disturbances among Hispanic/Latino, non-Hispanic (NH)-Asian, NH-American Indian/Alaska Native, NH-Black/African American, and NH-Other single and multiple races’ reporting satisfaction or dissatisfaction with life compared to non-Hispanic White adults reporting satisfaction with life, National Health Interview Survey, 2022, (N=24,314)

#### Various racial and ethnic groups compared to NH-White adults satisfied with life by age

Compared to NH-White adults who were satisfied with life, Hispanic/Latino adults aged 18-30 years with life satisfaction had 4% higher prevalence of infrequent insomnia symptoms (aPR=1.04, 95% CI: 0.99–1.09) and 8% lower prevalence of restorative sleep (aPR=0.92, 95% CI: 0.85–0.99), compared to NH-White adults who were satisfied with life. Whereas Hispanic/Latino adults in this age group reporting life dissatisfaction had 50% lower prevalence of infrequent insomnia symptoms (aPR=0.50, 95% CI: 0.29–0.86) and 69% lower prevalence of restorative sleep (aPR=0.31, 95% CI: 0.13–0.72).

Compared to NH-White adults who were satisfied with life, Hispanic/Latino adults aged 31 – 49 years with life satisfaction had a similar prevalence of recommended sleep duration (aPR=1.00, 95% CI: 0.94–1.05), whereas the same aged Hispanic/Latino adults reporting life dissatisfaction had 53% lower prevalence of recommended sleep duration (aPR=0.47, 95% CI: 0.27–0.84). Moreover, Hispanic/Latino adults aged 31 – 49 years with life satisfaction had 8% higher prevalence of infrequent insomnia symptoms (aPR=1.08, 95% CI: 1.05–1.12), while Hispanic/Latino adults in the same age range reporting life dissatisfaction had 26% lower prevalence of infrequent insomnia symptoms (aPR=0.74, 95% CI: 0.53–1.05) (Table 4).

NH-Black adults aged 31–49 years with life satisfaction had an 8% higher prevalence of infrequent insomnia symptoms (aPR=1.08, 95% CI: 1.03–1.12) and a comparable prevalence of restorative sleep (aPR=1.02, 95% CI: 0.94–1.12), compared to NH-White adults who were satisfied with life. In contrast, NH-Black adults reporting life dissatisfaction had substantially lower prevalence of both infrequent insomnia symptoms (aPR=0.49, 95% CI: 0.29–0.83) and restorative sleep (aPR=0.21, 95% CI: 0.08–0.58) (Table 4).

#### Racial and ethnic groups compared to NH-White adults satisfied with life by sex

Compared to NH-White adults who were satisfied with life, Hispanic/Latino men with life satisfaction had an 8% higher prevalence of infrequent insomnia symptoms (aPR=1.08, 95% CI: 1.05–1.11) and a similar prevalence of restorative sleep (aPR=0.99, 95% CI: 0.94–1.04). In contrast, Hispanic/Latino men reporting life dissatisfaction had substantially lower prevalence of both infrequent insomnia symptoms (35% lower; aPR=0.65, 95% CI: 0.46–0.92) and restorative sleep (52% lower; aPR=0.48, 95% CI: 0.28–0.83) (Table 4).

Among Hispanic/Latina women, women with life satisfaction had a 10% higher prevalence of infrequent insomnia symptoms (aPR=1.10, 95% CI: 1.06–1.13) and a comparable prevalence of restorative sleep (aPR=0.98, 95% CI: 0.92–1.03). However, life dissatisfaction was associated with lower prevalence of both outcomes: 24% lower for infrequent insomnia symptoms (aPR=0.76, 95% CI: 0.56–1.03) and 45% lower for restorative sleep (aPR=0.55, 95% CI: 0.34–0.88).

Similarly, NH-Black men with life satisfaction had a 15% lower prevalence of recommended sleep duration (aPR=0.85, 95% CI: 0.80–0.90), but a similar prevalence of restorative sleep (aPR=0.98, 95% CI: 0.92–1.04), compared to NH-White adults who were satisfied with life. NH-Black men reporting life dissatisfaction had a 42% lower prevalence of recommended sleep duration (aPR=0.58, 95% CI: 0.41–0.81) and a 45% lower prevalence of restorative sleep (aPR=0.55, 95% CI: 0.33–0.91).

## Discussion

This study is the first, to our knowledge, to leverage a large, nationally-representative sample of U.S. adults to investigate associations between life satisfaction and favorable sleep health – defined as meeting recommended sleep duration, reporting infrequent insomnia symptoms, and experiencing restorative sleep. Although life satisfaction varied by age, sex, and race along with ethnicity, these characteristics did not significantly modify associations; the relationship between life satisfaction and sleep outcomes remained consistent across demographic groups. However, differences in magnitude of associations were observed. For instance, the association between life satisfaction and restorative sleep was marginally stronger among younger adults and men, Similarly, associations between life satisfaction and higher prevalence of recommended sleep and infrequent insomnia symptoms were comparable across sexes. Race and ethnicity were observed to influence select aspects of the life satisfaction-sleep relationship. Stronger associations among Hispanic/Latino adults vs. NH-White and NH-Black, particularly for the link between life satisfaction and infrequent insomnia symptoms—possibly reflecting protective cultural factors such as robust social networks [38]. However, race and ethnicity did not significantly modify associations for recommended sleep duration and restorative sleep. Interrelated associations showed that NH-Asian, NH-Black, and NH-multiracial/Other adults satisfied with life satisfaction had a lower prevalence of recommended sleep compared to NH-White adults. Conversely, Hispanic/Latino, NH-Asian, and NH-Black adults with life satisfaction had higher prevalence of infrequent insomnia symptoms than their NH-White counterparts.

Plausible psychological mechanisms support the likely bidirectional nature of the relationship between life satisfaction and sleep. Although our study focused on life satisfaction as the exposure, findings are consistent with longitudinal evidence from Czech, German, and Swedish cohorts showing reciprocal relationships between sleep quality and life satisfaction during key life transitions [10, 13, 16, 18]. The stronger association with restorative sleep among young adults in our study similarly supports prior studies highlighting the critical role of sleep in early adulthood, a period of increasing self-reliance [11, 14, 15]. For older adults, prior studies assessing sleep in relation to life satisfaction in China, Finland, and Iran suggest retirement improves sleep and life satisfaction, with depression potentially mediating this link. These associations may be attenuated due to shifts in priorities or lifestyle factors such as retirement, which may alleviate stress and promote sleep health [17, 19–21, 39].

Furthermore, sex did not modify associations, but the magnitude of associations varied. Life satisfaction and a higher magnitude of prevalence of restorative sleep among men compared to women may reflect sex-based differences in hormonal changes, stress responses, or exposures to societal practices and policies. For example, differential access to socioeconomic resources and societal expectations may affect psychosocial pathways (e.g., increased stress) which in turn influences how men and women experience and report life satisfaction and sleep quality [25, 35, 40, 41].

Differences by race and ethnicity in income, housing, and neighborhood environments may contribute to poorer sleep even among individuals reporting high life satisfaction [42, 43]. However, in the current study, Hispanic/Latino adults with life satisfaction were associated with higher prevalence of recommended sleep, infrequent insomnia symptoms, and restorative sleep. Additionally, NH-Asian and NH-Black adults with life satisfaction had higher prevalence of infrequent insomnia symptoms which may also suggest the presence of adaptive coping mechanisms and culturally rooted resilience factors, such as strong familial and social support systems that buffer stress-related sleep disruptions [44–47]. Conversely, NH-Asian, NH-Black, and NH-multiracial/Other adults satisfied with life were associated with lower prevalence of recommended sleep compared to NH-White adults [26, 27]. Life satisfaction alone may be insufficient to promote sleep health in the presence of persistent sociocultural and environmental stressors, underscoring the need for theoretical frameworks that address both psychological and contextual determinants. For instance, bottom-up theories which conceptualize life satisfaction as the aggregation of satisfaction across domains (e.g., health, finances, relationships) or top-down theories that highlight the role of stable personality traits and cognitive appraisals [48–52].

This study has several limitations. First, the cross-sectional design precludes causal inference and limits our ability to determine the temporality between life satisfaction and sleep outcomes. Longitudinal studies can help establish directionality. Second, reliance on self-reported sleep introduces potential recall and social desirability bias, possibly resulting in misclassification [53]. Future studies using objective measures (e.g., actigraphy or polysomnography) could improve validity. Third, unmeasured or residual confounding may persist despite adjustment for key covariates; relevant lifestyle (e.g., religiosity, resiliency and social support) or psychosocial factors (e.g., trauma exposure) may not have been captured. Fourth, income was not assessed as a mediator in the model adjustment, although related variables (i.e., employment status, educational attainment) were included. Finally, data collection during the COVID-19 pandemic may have influenced both sleep and patterns of life satisfaction non-differentially but may limit generalizability to other periods of time.

Despite the limitations, this study has notable strengths. We used a nationally-representative sample of U.S. adults to determine the associations between life satisfaction and sleep health across sociodemographic groups, strengthening external validity to individuals living in the U.S. The large sample size enables robust statistical analyses and supports examination of interrelated associations in the life satisfaction-sleep relationship as well as test modification by key sociodemographic factors as well as potential mediators of the life satisfaction and sleep relationship.

In conclusion, life satisfaction was associated with recommended sleep duration, infrequent insomnia symptoms, and restorative sleep in a large, nationally representative sample of U.S. adults. These associations were largely consistent across age, sex, and race along with ethnicity, though magnitudes varied. For instance, higher prevalence of recommended sleep duration was observed among younger vs. older adults; restorative sleep was more prevalent among men vs. women; and infrequent insomnia symptoms were more common among Hispanic/Latino vs. NH-White and NH-Black adults. We also observed that NH-Asian, NH-Black, and NH-multiracial/other adults compared to NH-White adults satisfied with life had a lower prevalence of recommended sleep duration. Public health strategies that enhance life satisfaction—through psychological support, socioeconomic security, and cultural resilience—may also improve sleep. Future longitudinal studies incorporating objective sleep metrics and stress biomarkers, for instance, are warranted to elucidate causal pathways and inform targeted public health promotion strategies.

## Data Availability

The datasets generated during and/or analyzed during the current study are publicly available.

## Declarations

### Funding

This work was funded by the Intramural Program at the NIH, National Institute of Environmental Health Sciences (Z1AES103325 (CLJ)).

### Conflicts of interest/Competing interests

None declared Ethics approval: Not applicable; exempt

### Consent to participate

Not applicable

### Consent for publication

Not applicable

### Availability of data and material

The datasets generated during and/or analyzed during the current study are publicly available.

## Acknowledgements

The authors wish to thank the National Health Interview Survey (NHIS) participants. We also thank Dr. Kaitlyn Lawerence for her review of an earlier draft of the manuscript.

## Code availability

Stata, Version 15.1 (Statacorp, College Station, Texas).

## Author contributions

*Authors:* Bethany T. Ogbenna, Symielle A. Gaston, Wensu Zhou, Christopher Payne, W. B. Jackson II, Chandra L. Jackson

*Study concept:* CL. Jackson.

*Study design:* SA. Gaston, CL. Jackson, BT. Ogbenna

*Acquisition of data:* C. Payne.

*Statistical Analysis:* C. Payne.

*Interpretation of data:* BT. Ogbenna, SA. Gaston, C. Payne, WB. Jackson II, CL. Jackson.

*Drafting of the manuscript:* BT. Ogbenna,

*Critical revision of the manuscript for important intellectual content:* BT. Ogbenna, SA. Gaston, W. Zhou, C Payne, WB Jackson II, CL. Jackson.

*Administrative, technical, and material support:* CL. Jackson, SA. Gaston,

*Obtaining funding:* CL. Jackson.

*Study supervision:* CL. Jackson, WB. Jackson II.

*Final Approval:* BT. Ogbenna, SA. Gaston, W. Zhou, C. Payne, WB. Jackson II, CL. Jackson.

## Notes

### Competing Interest Statement

The authors have declared no competing interest.

