## Supplemental Figures and Tables for "Satisfaction with Life in relation to Sleep Health among a Nationally Representative Sample of U.S. Adults"

**Supplemental Figure 1. Flow chart of study population selection**

NHIS 2022

sample of adults

**(N=27,651)**

Final analytic sample adults

**N=25,090**

- Missing data
  - All sleep measures (n = 1,044)
  - Life satisfaction (n = 58)
  - Sex (n =3)
  - Race and ethnicity (n=0)
  - Age (n=56)
  - Potential confounders (n= 1,400)
- **Total excluded (n =2,561)**

**Excluded**

**Supplemental Table 1. Comparison of included and excluded study participants, National Health Interview Survey, 2022, (N=27,651)**

|  | Included n=25,090 (90.7%) | Excluded  n=2,561 (9.3%) | Chi-square or t-test p-value |
| --- | --- | --- | --- |
| Sociodemographic Characteristics |  |  |  |
| Age (years), mean (SE) | 48.1 (.17) | 66.6 (2.4) | <0.001 |
| Age group |  |  | 0.552 |
| 18-30 | 21.8 | 20.8 |  |
| 31-49 | 31.5 | 32.5 |  |
| ≥ 50 | 46.7 | 46.7 |  |
| Sex |  |  | <0.001 |
| Men | 49.1 | 44.5 |  |
| Women | 50.9 | 55.5 |  |
| Race/ethnicity |  |  | <0.001 |
| Hispanic/Latino | 16.9 | 20.4 |  |
| NH-American Indian/Alaska Native | 0.8 | 0.7 |  |
| NH-Asian | 6.1 | 5.3 |  |
| NH-Black/African American | 11.2 | 18.4 |  |
| NH-Multiracial or other group ^a^ | 2.0 | 2.0 |  |
| NH-White | 62.9 | 53.2 |  |
| Educational Attainment |  |  | <0.001 |
| ≤High School | 37.2 | 46.3 |  |
| Some College | 29.6 | 27.9 |  |
| ≥College | 33.2 | 25.8 |  |
| Employment status |  |  | <0.001 |
| Employed, currently working | 63.7 | 59.1 |  |
| Employed, not currently working | 0.6 | 0.5 |  |
| Not employed, Homemaker | 4.7 | 5.3 |  |
| Not employed, Going to school | 2.3 | 3.5 |  |
| Not employed, Retired | 18.9 | 18.9 |  |
| Not employed, not able to work | 6.0 | 9.4 |  |
| Not employed, looking for work | 2.2 | 2.1 |  |
| Not employed, Other | 1.6 | 1.2 |  |
| Marital status |  |  | <0.001 |
| Divorced/widowed | 17.2 | 19.3 |  |
| Single/no live-in partner | 23.6 | 28.8 |  |
| Married/living with partner/co-habiting | 59.2 | 51.9 |  |
| Region of residence |  |  | <0.001 |
| Northeast | 17.2 | 21.2 |  |
| Midwest | 21.0 | 17.4 |  |
| South | 37.9 | 39.7 |  |
| West | 23.8 | 21.6 |  |
| Health Behaviors |  |  |  |
| Smoking status |  |  | 0.161 |
| Never/quit >12 months prior | 87.3 | 85.7 |  |
| Former/quit≤12 months ago | 1.2 | 1.1 |  |
| Current | 11.5 | 13.2 |  |
| Alcohol consumption |  |  | <0.001 |
| Current (≥1 drink past year) | 69.9 | 61.3 |  |
| Former (no drinks past year) | 17.0 | 18.5 |  |
| Lifetime abstinence (<12 drinks in life) | 13.0 | 20.2 |  |
| Leisure-time physical activity ^b^ |  |  | <0.001 |
| Inactive | 26.5 | 38.0 |  |
| Insufficiently active | 25.6 | 24.1 |  |
| Sufficiently active | 47.8 | 37.9 |  |
| Usual sleep duration |  |  | 0.045 |
| Short (<7 hours) | 30.3 | 27.2 |  |
| Recommended (≥7 hours) | 69.7 | 72.8 |  |
| Infrequent Insomnia symptoms (yes) ^c^ | 23.3 | 22.7 | 0.691 |
| Restorative sleep (yes) ^d^ | 43.0 | 45.1 | 0.163 |
| Clinical Characteristics |  |  |  |
| Ever had depression (yes) ^e^ | 18.5 | 16.4 | 0.028 |
| Body mass index category |  |  | 0.391 |
| Underweight (<18.5 kg/m^2^) | 1.7 | 1.8 |  |
| Recommended (18.5-<25 kg/m^2^) | 31.2 | 32.1 |  |
| Overweight (25-<30 kg/m^2^) | 33.7 | 35.0 |  |
| Obesity (≥30 kg/m^2^) | 33.4 | 31.1 |  |
| General health status |  |  | <0.001 |
| Fair/poor | 13.9 | 19.8 |  |
| Good/very good/excellent | 86.1 | 80.2 |  |
| Abbreviations: SE (standard error), NH (non-Hispanic) | | | |
| Note: Data are presented as column percentages or means and standard errors. Percentages may not sum to 100 due to missing rounding. All estimates are weighted for the survey's complex sampling design. All estimates are age-standardized to the U.S. 2020 population, except for age. | | | |
| ^a^ NH-Other single and multiple races and ‘Other group’ is defined as persons identifying with racial groups not explicitly listed in the standard categories.  ^b^ Leisure-time physical activity was defined using the 2018 Health and Human Services Physical Activity guidelines which state “recommend that adults complete at least 150 minutes to 300 minutes of moderate-intensity activity, or 75 minutes to 150 minutes of vigorous-intensity aerobic activity per week, as well as moderate or greater intensity muscle strengthening activities on two or more days a week."  ^c^ Infrequent Insomnia symptoms (yes) defined as difficulty falling or staying asleep most days/every day. | | | |
| ^d^ Restorative sleep (yes) defined as never/some days waking up feeling rested in the past 30 days. | | | |
| ^e^ Depression was defined by the 2019 Field Representative's Manual as a major depressive disorder or as clinical depression that is a common but serious mood disorder. It causes severe symptoms that affect how you feel, think, and handle daily activities, such as sleeping, eating, or working.” | | | |

**Supplemental Table 2. Study population characteristics, overall and by age, National Health Interview Survey, 2022, (N=25,090)**

|  | Total  n=25,090 (100%) | | | 18-30 years  n=3,639 (14.5%) | | | 31-49 years  n=7,198 (28.7%) | | | ≥ 50 years  n=14,253 (56.8%) | | |
| --- | --- | --- | --- | --- | --- | --- | --- | --- | --- | --- | --- | --- |
|  |  | Life satisfaction ^a^ | |  | Life satisfaction ^a^ | |  | Life satisfaction ^a^ | |  | Life satisfaction ^a^ | |
|  | All  n=25,090 (100%) | Yes n=23,997 (95.6%) | No  n=1,093 (4.4%) | All  n=3,639 (100%) | Yes  n=3,515 (96.6%) | No  n=124 (3.4%) | All  n=7,198 (100%) | Yes  n=6,941 (96.4%) | No  n=257 (3.6%) | All  n=14,253 (100%) | Yes n=13,541 (95.0%) | No  n=712 (5.0%) |
| Sociodemographic Characteristics |  | | |  | | |  | | |  | | |
| Age (years), mean (SE) | 48.1(.17) | 48.0(.17) | 50.5(.75) | 24.1(.07) | 24.1(.07) | 24.1(.37) | 39.7(.07) | 39.7(.08) | 39.9(.39) | 64.9(.10) | 64.9(.11) | 65.4(.47) |
| Race and ethnicity |  | | |  | | |  | | |  | | |
| Hispanic/Latino | 16.9 | 17.1 | 13.6 | 22.6 | 22.7 | 19.7 | 20.0 | 20.2 | 13.7 | 12.2 | 12.3 | 10.5 |
| NH-American Indian/Alaska Native | 0.8 | 0.8 | 1.0 | 1.2 | 1.2 | 2.4 | 0.8 | 0.8 | 0.2 | 0.6 | 0.6 | 0.7 |
| NH-Asian | 6.1 | 6.3 | 2.7 | 5.7 | 5.9 | 1.0 | 7.7 | 7.8 | 3.5 | 5.3 | 5.4 | 3.1 |
| NH-Black/African American | 11.2 | 11.1 | 13.5 | 12.5 | 12.2 | 19.2 | 12.0 | 11.9 | 14.0 | 10.0 | 10.0 | 10.4 |
| NH-Multiracial or other group ^b^ | 2.0 | 2.0 | 3.4 | 3.7 | 3.7 | 4.3 | 2.1 | 2.0 | 5.0 | 1.2 | 1.1 | 1.9 |
| NH-White | 62.9 | 62.8 | 65.9 | 54.3 | 54.4 | 53.3 | 57.5 | 57.2 | 63.6 | 70.8 | 70.6 | 73.5 |
| Sex |  | | |  | | |  | | |  | | |
| Men | 49.1 | 49.0 | 51.3 | 50.6 | 50.2 | 63.2 | 49.8 | 49.8 | 49.9 | 47.9 | 48.0 | 46.2 |
| Women | 50.9 | 51.0 | 48.7 | 49.4 | 49.8 | 36.8 | 50.2 | 50.2 | 50.1 | 52.1 | 52.0 | 53.8 |
| Educational Attainment |  | | |  | | |  | | |  | | |
| ≤ High School | 37.2 | 36.8 | 47.3 | 40.9 | 40.6 | 50.2 | 31.2 | 30.7 | 45.0 | 39.5 | 39.2 | 47.2 |
| Some college | 29.6 | 29.5 | 33.6 | 33.7 | 33.6 | 36.2 | 28.2 | 28.0 | 34.1 | 28.7 | 28.5 | 31.8 |
| ≥ College | 33.2 | 33.8 | 19.2 | 25.4 | 25.8 | 13.7 | 40.6 | 41.3 | 20.9 | 31.8 | 32.3 | 21.0 |
| Employment status |  | | |  | | |  | | |  | | |
| Employed, currently working | 63.7 | 64.6 | 41.0 | 75.5 | 76.0 | 62.4 | 82.1 | 83.1 | 52.7 | 45.2 | 46.3 | 23.1 |
| Employed, not currently working | 0.6 | 0.6 | 0.9 | 0.5 | 0.5 | 0.8 | 0.7 | 0.6 | 1.1 | 0.6 | 0.6 | 0.7 |
| Not employed, Homemaker | 4.7 | 4.7 | 4.7 | 5.1 | 5.0 | 7.5 | 7.8 | 7.8 | 7.1 | 2.5 | 2.5 | 1.8 |
| Not employed, Going to school | 2.3 | 2.4 | 1.6 | 9.8 | 9.9 | 7.0 | 0.5 | 0.5 | 0.0 | 0.1 | 0.1 | 0.0 |
| Not employed, Retired | 18.9 | 19.0 | 16.8 | 0.0 | 0.0 | 0.0 | 0.3 | 0.3 | 0.0 | 40.9 | 41.1 | 36.3 |
| Not employed, not able to work | 6.0 | 5.1 | 25.0 | 2.4 | 2.4 | 3.5 | 4.7 | 3.9 | 28.1 | 8.4 | 7.2 | 33.0 |
| Not employed, looking for work | 2.2 | 2.0 | 6.3 | 4.5 | 4.1 | 12.6 | 2.1 | 1.9 | 5.4 | 1.2 | 1.1 | 3.7 |
| Not employed, Other | 1.6 | 1.5 | 3.8 | 2.3 | 2.1 | 6.2 | 1.9 | 1.7 | 5.6 | 1.1 | 1.1 | 1.4 |
| Marital status |  | | |  | | |  | | |  | | |
| Divorced/widowed | 17.2 | 16.7 | 28.0 | 1.9 | 1.8 | 4.2 | 11.0 | 10.7 | 21.5 | 28.7 | 27.9 | 43.9 |
| Single/no live-in partner | 23.6 | 23.1 | 37.4 | 65.9 | 65.4 | 77.7 | 17.9 | 17.1 | 41.7 | 7.7 | 7.3 | 14.7 |
| Married/living with partner/co-habited | 59.2 | 60.2 | 34.6 | 32.2 | 32.7 | 18.1 | 71.1 | 72.2 | 36.8 | 63.7 | 64.8 | 41.4 |
| Region of residence |  | | |  | | |  | | |  | | |
| Northeast | 17.2 | 17.2 | 17.8 | 17.6 | 17.4 | 23.0 | 16.1 | 16.1 | 14.0 | 17.8 | 17.8 | 17.6 |
| Midwest | 21.0 | 21.0 | 22.1 | 21.1 | 21.2 | 20.9 | 20.2 | 20.1 | 23.0 | 21.5 | 21.5 | 22.0 |
| South | 37.9 | 38.0 | 35.4 | 37.9 | 38.2 | 29.4 | 37.9 | 37.9 | 38.5 | 37.9 | 38.0 | 36.8 |
| West | 23.8 | 23.8 | 24.7 | 23.4 | 23.3 | 26.8 | 25.8 | 25.9 | 24.6 | 22.7 | 22.7 | 23.6 |
| **Health Behaviors** |  | | |  | | |  | | |  | | |
| Smoking status |  | | |  | | |  | | |  | | |
| Never/quit>12 months prior | 87.3 | 87.8 | 73.9 | 91.0 | 91.4 | 81.9 | 84.8 | 85.7 | 59.7 | 87.2 | 87.6 | 78.9 |
| Former/quit≤12 months ago | 1.2 | 1.2 | 2.4 | 1.8 | 1.7 | 3.4 | 1.4 | 1.4 | 1.0 | 0.8 | 0.7 | 2.8 |
| Current | 11.5 | 11.1 | 23.7 | 7.2 | 6.9 | 14.8 | 13.9 | 13.0 | 39.2 | 12.0 | 11.6 | 18.3 |
| Alcohol consumption |  | | |  | | |  | | |  | | |
| Current (≥1 drink past year) | 69.9 | 70.2 | 64.4 | 72.4 | 72.2 | 77.5 | 76.9 | 77.1 | 71.9 | 64.0 | 64.5 | 53.2 |
| Former (no drinks past year) | 17.0 | 16.7 | 23.8 | 8.3 | 8.2 | 9.2 | 12.2 | 12.0 | 17.6 | 24.5 | 23.9 | 35.1 |
| Lifetime abstinence (<12 drinks in life) | 13.0 | 13.1 | 11.7 | 19.3 | 19.6 | 13.3 | 10.9 | 10.9 | 10.5 | 11.6 | 11.6 | 11.7 |
| Leisure-time physical activity ^c^ |  | | |  | | |  | | |  | | |
| Inactive | 26.5 | 25.8 | 44.3 | 19.0 | 18.9 | 20.9 | 22.7 | 22.0 | 43.5 | 32.7 | 31.6 | 56.4 |
| Insufficiently active | 25.6 | 25.7 | 23.4 | 22.9 | 22.9 | 25.5 | 27.6 | 27.8 | 22.5 | 25.5 | 25.6 | 22.9 |
| Sufficiently active | 47.8 | 48.5 | 32.3 | 58.1 | 58.2 | 53.6 | 49.6 | 50.2 | 34.0 | 41.8 | 42.8 | 20.8 |
| Usual sleep duration |  | | |  | | |  | | |  | | |
| Short (<7 hours) | 30.3 | 29.7 | 45.4 | 26.7 | 26.0 | 45.7 | 34.3 | 33.8 | 50.0 | 29.1 | 28.5 | 41.8 |
| Recommended (≥7 hours) | 69.7 | 70.3 | 54.6 | 73.3 | 74.0 | 54.3 | 65.7 | 66.2 | 50.0 | 70.9 | 71.5 | 58.2 |
| Infrequent insomnia symptoms (yes) ^d^ | 76.7 | 77.8 | 49.6 | 77.8 | 78.8 | 52.6 | 78.8 | 79.8 | 48.1 | 74.8 | 76.1 | 49.6 |
| Restorative sleep (yes) ^e^ | 57.0 | 58.3 | 25.7 | 54.6 | 55.9 | 21.6 | 51.9 | 53.0 | 21.8 | 61.6 | 63.1 | 30.5 |
| **Clinical Characteristics** |  | | |  | | |  | | |  | | |
| Ever had depression (yes) ^f^ | 18.5 | 16.9 | 55.2 | 21.0 | 19.7 | 54.0 | 18.0 | 16.7 | 56.1 | 17.5 | 15.7 | 55.2 |
| Body mass index category |  | | |  | | |  | | |  | | |
| Underweight (<18.5 kg/m^2^) | 1.7 | 1.6 | 2.8 | 3.5 | 3.5 | 3.5 | 0.9 | 0.9 | 2.3 | 1.3 | 1.2 | 2.8 |
| Recommended (18.5-<25 kg/m^2^) | 31.2 | 31.3 | 28.2 | 42.4 | 42.5 | 38.9 | 28.0 | 28.2 | 24.2 | 28.2 | 28.3 | 25.4 |
| Overweight (25-<30 kg/m^2^) | 33.7 | 34.0 | 27.0 | 28.3 | 28.5 | 24.4 | 34.0 | 34.4 | 20.7 | 36.1 | 36.3 | 32.0 |
| Obesity (≥30 kg/m^2^) | 33.4 | 33.0 | 42.0 | 25.7 | 25.4 | 33.1 | 37.1 | 36.5 | 52.9 | 34.4 | 34.1 | 39.8 |
| General health status |  | | |  | | |  | | |  | | |
| Fair/poor | 13.9 | 12.3 | 51.8 | 6.2 | 5.2 | 32.1 | 9.8 | 8.5 | 48.7 | 20.4 | 18.3 | 63.4 |
| Good/very good/excellent | 86.1 | 87.7 | 48.2 | 93.8 | 94.8 | 67.9 | 90.2 | 91.5 | 51.3 | 79.6 | 81.7 | 36.6 |
| Abbreviations: SE (standard error), NH (non-Hispanic) | | | | | | | | | | | | |
| Note: Data are presented as column percentages or means and standard errors. Percentages may not sum to 100 due to missing rounding. All estimates are weighted for the survey's complex sampling design. All estimates are age-standardized to theU.S.2020 population, except for age. | | | | | | | | | | | | |
| ^a^ Life satisfaction was ascertained based on the questions, 'In general, how satisfied are you with your life? Are you very satisfied, satisfied, dissatisfied, or very dissatisfied?'. Life satisfaction was dichotomized as yes (a response of 'satisfied' or 'very satisfied') vs. no (a response of 'very dissatisfied' or 'dissatisfied'). | | | | | | | | | | | | |
| ^b^ NH-Other single and multiple races and ‘Other group’ is defined as persons identifying with racial groups not explicitly listed in the standard categories.  ^c^ Leisure-time physical activity was defined using the 2018 Health and Human Services Physical Activity guidelines which state “recommend that adults complete at least 150 minutes to 300 minutes of moderate-intensity activity, or 75 minutes to 150 minutes of vigorous-intensity aerobic activity per week, as well as moderate or greater intensity muscle strengthening activities on two or more days a week."  ^d^ Infrequent Insomnia symptoms (yes) defined as difficulty falling or staying asleep most days/every day. | | | | | | | | | | | | |
| ^e^ Restorative sleep (yes) defined as never/some days waking up feeling rested in the past 30 days. | | | | | | | | | | | | |
| ^f^ Depression was defined by the 2019 Field Representative's Manual as a major depressive disorder or as clinical depression that is a common but serious mood disorder. It causes severe symptoms that affect how you feel, think, and handle daily activities, such as sleeping, eating, or working.” | | | | | | | | | | | | |

**Supplemental Table 3. Study population characteristics, overall and by race and ethnicity, National Health Interview Survey, 2022, (N=25,090)**

|  | Total  n=25,090 (100%) | | | Hispanic/Latino n=3,511 (14.0%) | | | NH-American Indian/Alaska Native n=172 (0.7%) | | | NH-Asian  n=1,531 (6.1%) | | | NH-Black/African American  n=2,664 (10.6%) | | | NH-Multiracial or other group ^a^  n=456 (1.8%) | | | NH-White  n=16,756 (66.8%) | | |
| --- | --- | --- | --- | --- | --- | --- | --- | --- | --- | --- | --- | --- | --- | --- | --- | --- | --- | --- | --- | --- | --- |
|  |  | Life satisfaction ^b^ | |  | Life satisfaction ^b^ | |  | Life satisfaction ^b^ | |  | Life satisfaction ^b^ | |  | Life satisfaction ^b^ | |  | Life satisfaction ^b^ | |  | Life satisfaction ^b^ | |
|  | All n=25,090 (100%) | Yes n=23,997 (95.6%) | No n=1,093 (4.4%) | All n=3,511 (100%) | Yes n=3,389 (96.5%) | No n=122 (3.5%) | All n=172 (100%) | Yes n=160 (93.0%) | No  n=12 (7.0%) | All n=1,531 (100%) | Yes n=1,500 (98.0%) | No  n=31 (2.0%) | All n=2,664 (100%) | Yes n=2,540 (95.3%) | No n=124 (4.7%) | All n=456 (100%) | Yes n=428 (93.9%) | No  n=28 (6.1%) | All n=16,756 (100%) | Yes n=15,980 (95.4%) | No n=776 (4.6%) |
| Sociodemographic Characteristics |  | | |  | | |  | | |  | | |  | | |  | | |  | | |
| Age (years), mean (SE) | 48.1(.17) | 48.0(.17) | 50.5(.75) | 42.6(.33) | 42.5(.33) | 46.4(2.3) | 43.5(1.5) | 43.7(1.5) | 39.0(6.1) | 46.3(.56) | 46.2(.57) | 53.0(2.7) | 46.0(.43) | 46.0(.44) | 44.7(2.0) | 39.0(.93) | 38.8(.96) | 41.9(3.1) | 50.4(.21) | 50.3(.21) | 52.9(.90) |
| 18-30 years | 21.8 | 21.9 | 20.0 | 29.1 | 29.1 | 29.9 | 33.7 | 32.8 | 51.8 | 20.3 | 20.5 | 7.5 | 24.4 | 24.1 | 29.4 | 40.2 | 41.1 | 26.9 | 18.8 | 18.9 | 16.0 |
| 31-49 years | 31.5 | 31.7 | 26.1 | 37.2 | 37.5 | 27.2 | 32.5 | 33.8 | 6.8 | 39.4 | 39.5 | 32.7 | 33.6 | 33.9 | 27.9 | 32.9 | 32.4 | 40.5 | 28.7 | 28.9 | 24.8 |
| ≥ 50 years | 46.7 | 46.4 | 53.9 | 33.7 | 33.4 | 42.9 | 33.8 | 33.4 | 41.4 | 40.3 | 40.0 | 59.8 | 42.0 | 42.0 | 42.6 | 26.9 | 26.5 | 32.5 | 52.5 | 52.2 | 59.2 |
| Sex |  | | |  | | |  | | |  | | |  | | |  | | |  | | |
| Men | 49.1 | 49.0 | 51.3 | 49.0 | 48.9 | 49.6 | 38.9 | 37.5 | 69.2 | 45.6 | 45.6 | 53.2 | 44.4 | 44.1 | 50.8 | 52.4 | 51.7 | 61.9 | 50.5 | 50.5 | 50.9 |
| Women | 50.9 | 51.0 | 48.7 | 51.0 | 51.1 | 50.4 | 61.1 | 62.5 | 30.8 | 54.4 | 54.4 | 46.8 | 55.6 | 55.9 | 49.2 | 47.6 | 48.3 | 38.1 | 49.5 | 49.5 | 49.1 |
| Educational Attainment |  | | |  | | |  | | |  | | |  | | |  | | |  | | |
| ≤ High School | 37.2 | 36.8 | 47.3 | 57.1 | 56.8 | 65.1 | 49.1 | 49.5 | 41.5 | 29.9 | 29.1 | 70.3 | 45.5 | 45.6 | 42.3 | 36.2 | 35.2 | 47.2 | 31.4 | 30.8 | 44.4 |
| Some college | 29.6 | 29.5 | 33.6 | 26.7 | 26.7 | 24.0 | 36.3 | 36.3 | 34.5 | 17.4 | 17.6 | 12.7 | 31.7 | 31.1 | 43.0 | 38.0 | 38.4 | 33.3 | 30.5 | 30.3 | 33.6 |
| ≥ College | 33.2 | 33.8 | 19.2 | 16.3 | 16.5 | 10.9 | 14.6 | 14.1 | 24.0 | 52.6 | 53.3 | 17.0 | 22.9 | 23.3 | 14.7 | 25.9 | 26.5 | 19.5 | 38.1 | 38.8 | 22.0 |
| Employment status |  | | |  | | |  | | |  | | |  | | |  | | |  | | |
| Employed, currently working | 63.7 | 64.6 | 41.0 | 64.4 | 64.9 | 45.4 | 49.5 | 51.1 | 13.9 | 62.5 | 63.1 | 31.9 | 61.2 | 62.2 | 40.7 | 62.8 | 65.0 | 31.9 | 64.6 | 65.7 | 41.0 |
| Employed, not currently working | 0.6 | 0.6 | 0.9 | 0.5 | 0.5 | 1.1 | 2.4 | 2.5 | 0.0 | 0.6 | 0.6 | 0.0 | 0.6 | 0.6 | 1.5 | 0.5 | 0.2 | 5.9 | 0.6 | 0.6 | 0.3 |
| Not employed, Homemaker | 4.7 | 4.7 | 4.7 | 7.1 | 7.2 | 2.5 | 8.7 | 9.1 | 0.0 | 6.9 | 6.8 | 8.4 | 2.7 | 2.6 | 5.5 | 4.9 | 5.2 | 0.0 | 4.2 | 4.2 | 5.0 |
| Not employed, Going to school | 2.3 | 2.4 | 1.6 | 2.0 | 2.1 | 0.0 | 2.2 | 2.3 | 0.0 | 5.1 | 5.1 | 0.0 | 2.6 | 2.5 | 3.3 | 2.6 | 2.7 | 2.2 | 2.1 | 2.1 | 1.7 |
| Not employed, Retired | 18.9 | 19.0 | 16.8 | 15.5 | 15.6 | 14.0 | 18.8 | 19.1 | 11.1 | 17.1 | 17.1 | 19.3 | 18.3 | 18.6 | 12.0 | 17.1 | 17.0 | 19.5 | 19.7 | 19.8 | 17.8 |
| Not employed, not able to work | 6.0 | 5.1 | 25.0 | 5.7 | 4.8 | 30.8 | 11.4 | 10.2 | 37.6 | 2.9 | 2.4 | 26.7 | 9.2 | 8.5 | 23.1 | 9.9 | 7.9 | 37.2 | 5.8 | 4.9 | 24.4 |
| Not employed, looking for work | 2.2 | 2.0 | 6.3 | 2.9 | 2.8 | 5.2 | 4.5 | 3.1 | 31.9 | 2.9 | 2.8 | 5.3 | 3.5 | 3.5 | 5.2 | 0.8 | 0.5 | 3.4 | 1.7 | 1.5 | 6.4 |
| Not employed, Other | 1.6 | 1.5 | 3.8 | 2.0 | 2.0 | 1.1 | 2.7 | 2.5 | 5.5 | 2.1 | 2.0 | 8.4 | 1.9 | 1.5 | 8.8 | 1.4 | 1.5 | 0.0 | 1.4 | 3.5 | 5.2 |
| Marital status |  | | |  | | |  | | |  | | |  | | |  | | |  | | |
| Divorced/widowed | 17.2 | 16.7 | 28.0 | 18.9 | 18.8 | 23.6 | 24.7 | 24.9 | 19.1 | 12.0 | 11.7 | 23.6 | 23.4 | 23.2 | 27.7 | 22.4 | 22.5 | 22.1 | 16.1 | 15.5 | 29.5 |
| Single/no live-in partner | 23.6 | 23.1 | 37.4 | 23.6 | 23.1 | 38.2 | 29.5 | 27.9 | 62.4 | 21.0 | 20.7 | 35.3 | 37.0 | 36.6 | 45.9 | 28.6 | 27.9 | 37.2 | 20.9 | 20.3 | 34.8 |
| Married/living with partner/co-habited | 59.2 | 60.2 | 34.6 | 57.5 | 58.1 | 38.3 | 45.8 | 47.2 | 18.6 | 67.0 | 67.6 | 41.1 | 39.6 | 40.2 | 26.4 | 49.0 | 49.6 | 40.7 | 63.0 | 64.2 | 35.7 |
| Region of residence |  | | |  | | |  | | |  | | |  | | |  | | |  | | |
| Northeast | 17.2 | 17.2 | 17.8 | 13.7 | 13.6 | 17.6 | 5.6 | 5.5 | 5.5 | 21.2 | 21.3 | 12.3 | 14.8 | 14.5 | 21.0 | 8.8 | 8.8 | 8.5 | 18.8 | 18.8 | 18.3 |
| Midwest | 21.0 | 21.0 | 22.1 | 8.1 | 8.1 | 9.3 | 14.3 | 14.3 | 15.1 | 13.4 | 13.3 | 23.4 | 14.2 | 14.0 | 17.6 | 16.9 | 16.8 | 19.3 | 26.6 | 26.6 | 25.4 |
| South | 37.9 | 38.0 | 35.4 | 39.1 | 39.0 | 37.2 | 26.9 | 27.0 | 26.9 | 25.1 | 25.1 | 22.4 | 62.2 | 62.7 | 51.8 | 32.2 | 32.8 | 23.6 | 34.7 | 34.8 | 32.7 |
| West | 23.8 | 23.8 | 24.7 | 39.1 | 39.4 | 35.9 | 53.2 | 53.2 | 52.6 | 40.3 | 40.3 | 41.8 | 8.8 | 8.8 | 9.6 | 42.1 | 41.7 | 48.7 | 20.0 | 19.8 | 23.5 |
| **Health Behaviors** |  | | |  | | |  | | |  | | |  | | |  | | |  | | |
| Smoking status |  | | |  | | |  | | |  | | |  | | |  | | |  | | |
| Never/quit>12 months prior | 87.3 | 87.8 | 73.9 | 90.7 | 91.0 | 82.9 | 75.7 | 74.8 | 93.5 | 94.5 | 94.7 | 83.1 | 84.2 | 84.7 | 74.0 | 85.2 | 86.1 | 72.3 | 86.2 | 86.9 | 70.1 |
| Former/quit≤12 months ago | 1.2 | 1.2 | 2.4 | 1.1 | 1.0 | 4.0 | 5.9 | 6.2 | 0.0 | 0.7 | 0.8 | 0.0 | 1.4 | 1.4 | 1.2 | 2.8 | 2.6 | 4.5 | 1.2 | 1.1 | 2.5 |
| Current | 11.5 | 11.1 | 23.7 | 8.3 | 8.0 | 13.1 | 18.5 | 19.1 | 6.5 | 4.8 | 4.6 | 16.9 | 14.4 | 13.9 | 24.8 | 12.1 | 11.3 | 23.1 | 12.6 | 12.0 | 27.4 |
| Alcohol consumption |  | | |  | | |  | | |  | | |  | | |  | | |  | | |
| Current (≥1 drink past year) | 69.9 | 70.2 | 64.4 | 62.6 | 62.9 | 53.7 | 50.3 | 51.0 | 36.9 | 52.6 | 52.8 | 54.3 | 63.3 | 63.1 | 68.2 | 69.6 | 69.4 | 74.4 | 74.7 | 75.1 | 66.7 |
| Former (no drinks past year) | 17.0 | 16.7 | 23.8 | 17.9 | 17.7 | 24.1 | 33.8 | 32.3 | 63.1 | 13.3 | 13.4 | 10.9 | 18.9 | 18.9 | 19.3 | 19.3 | 19.2 | 18.3 | 16.7 | 16.3 | 24.6 |
| Lifetime abstinence (<12 drinks in life) | 13.0 | 13.1 | 11.7 | 19.6 | 19.4 | 22.1 | 15.9 | 16.7 | 0.0 | 34.1 | 33.9 | 34.8 | 17.7 | 18.0 | 12.5 | 11.1 | 11.4 | 7.3 | 8.6 | 8.6 | 8.7 |
| Leisure-time physical activity ^c^ |  | | |  | | |  | | |  | | |  | | |  | | |  | | |
| Inactive | 26.5 | 25.8 | 44.3 | 35.3 | 34.7 | 51.8 | 30.3 | 29.1 | 57.7 | 24.0 | 23.5 | 37.6 | 31.7 | 31.2 | 41.8 | 29.2 | 28.3 | 43.6 | 23.5 | 22.6 | 43.6 |
| Insufficiently active | 25.6 | 25.7 | 23.4 | 24.9 | 25.1 | 18.4 | 29.0 | 29.9 | 11.0 | 28.9 | 28.9 | 30.8 | 26.3 | 26.8 | 16.9 | 23.2 | 23.4 | 20.2 | 25.3 | 25.2 | 26.2 |
| Sufficiently active | 47.8 | 48.5 | 32.3 | 39.8 | 40.2 | 29.7 | 40.7 | 41.0 | 31.4 | 47.1 | 47.6 | 31.6 | 42.0 | 42.0 | 41.3 | 47.6 | 48.4 | 36.2 | 51.3 | 52.2 | 30.2 |
| Usual sleep duration |  | | |  | | |  | | |  | | |  | | |  | | |  | | |
| Short (<7 hours) | 30.3 | 29.7 | 45.4 | 31.1 | 30.4 | 52.2 | 37.0 | 35.5 | 65.9 | 27.6 | 27.5 | 43.1 | 39.3 | 38.4 | 57.9 | 38.1 | 37.3 | 51.5 | 28.4 | 27.9 | 40.2 |
| Recommended (≥7 hours) | 69.7 | 70.3 | 54.6 | 68.9 | 69.6 | 47.8 | 63.0 | 64.5 | 34.1 | 72.4 | 72.5 | 56.9 | 60.7 | 61.6 | 42.1 | 61.9 | 62.7 | 48.5 | 71.6 | 72.1 | 59.8 |
| Infrequent Insomnia symptoms (yes) ^d^ | 76.7 | 77.8 | 49.6 | 81.7 | 83.1 | 42.0 | 66.4 | 67.8 | 37.5 | 87.3 | 87.7 | 62.3 | 79.4 | 80.5 | 57.4 | 72.3 | 74.2 | 45.4 | 74.3 | 75.3 | 50.4 |
| Restorative sleep (yes) ^e^ | 57.0 | 58.3 | 25.7 | 55.8 | 57.0 | 22.1 | 55.4 | 56.1 | 39.9 | 63.5 | 64.0 | 32.7 | 54.4 | 55.6 | 29.3 | 53.5 | 54.3 | 38.2 | 57.0 | 58.5 | 24.5 |
| **Clinical Characteristics** |  | | |  | | |  | | |  | | |  | | |  | | |  | | |
| Ever had depression (yes) ^f^ | 18.5 | 16.9 | 55.2 | 14.0 | 12.6 | 55.9 | 23.5 | 22.3 | 49.3 | 7.4 | 6.7 | 46.5 | 14.3 | 12.7 | 46.5 | 18.5 | 17.0 | 40.7 | 21.7 | 20.1 | 58.8 |
| Body mass index category |  | | |  | | |  | | |  | | |  | | |  | | |  | | |
| Underweight (<18.5 kg/m^2^) | 1.7 | 1.6 | 2.8 | 1.0 | 0.9 | 2.1 | 2.5 | 2.7 | 0.0 | 3.5 | 3.6 | 0.0 | 1.3 | 1.4 | 0.0 | 2.5 | 2.7 | 0.0 | 1.7 | 1.6 | 3.9 |
| Recommended (18.5-<25 kg/m^2^) | 31.2 | 31.3 | 28.2 | 23.7 | 23.8 | 19.2 | 29.1 | 27.6 | 56.2 | 54.1 | 54.4 | 27.6 | 23.4 | 23.1 | 30.0 | 34.5 | 34.9 | 26.0 | 32.6 | 32.8 | 28.9 |
| Overweight (25-<30 kg/m^2^) | 33.7 | 34.0 | 27.0 | 37.5 | 37.7 | 33.5 | 31.4 | 31.8 | 24.4 | 30.5 | 30.4 | 43.7 | 32.2 | 32.7 | 22.3 | 28.4 | 27.9 | 38.4 | 33.5 | 33.8 | 26.3 |
| Obesity (≥30 kg/m^2^) | 33.4 | 33.0 | 42.0 | 37.7 | 37.5 | 45.2 | 37.0 | 37.9 | 19.4 | 11.9 | 11.5 | 28.7 | 43.0 | 42.8 | 47.7 | 34.6 | 34.5 | 35.6 | 32.2 | 31.8 | 40.9 |
| General health status |  | | |  | | |  | | |  | | |  | | |  | | |  | | |
| Fair/poor | 13.9 | 12.3 | 51.8 | 18.3 | 17.0 | 58.2 | 20.9 | 19.2 | 58.1 | 11.0 | 10.2 | 52.0 | 19.4 | 17.6 | 54.5 | 18.1 | 16.1 | 48.4 | 12.4 | 10.6 | 51.1 |
| Good/very good/excellent | 86.1 | 87.7 | 48.2 | 81.7 | 83.0 | 41.8 | 79.1 | 80.8 | 41.9 | 89.0 | 89.8 | 48.0 | 80.6 | 82.4 | 45.5 | 81.9 | 83.9 | 51.6 | 87.6 | 89.4 | 48.9 |
| Abbreviations: SE (standard error), NH (non-Hispanic) | | | | | | | | | | | | | | | | | | | | | |
| Note: Data are presented as column percentages or means and standard errors. Percentages may not sum to 100 due to missing rounding. All estimates are weighted for the survey's complex sampling design. All estimates are age-standardized to theU.S.2020 population, except for age. | | | | | | | | | | | | | | | | | | | | | |
| ^a^ NH-Other single and multiple races and ‘Other group’ is defined as persons identifying with racial groups not explicitly listed in the standard categories.  ^b^ Life satisfaction was ascertained based on the questions, 'In general, how satisfied are you with your life? Are you very satisfied, satisfied, dissatisfied, or very dissatisfied?'. Life satisfaction was dichotomized as yes (a response of 'satisfied' or 'very satisfied') vs. no (a response of 'very dissatisfied' or 'dissatisfied'). | | | | | | | | | | | | | | | | | | | | | |
| ^c^ Leisure-time physical activity was defined using the 2018 Health and Human Services Physical Activity guidelines which state “recommend that adults complete at least 150 minutes to 300 minutes of moderate-intensity activity, or 75 minutes to 150 minutes of vigorous-intensity aerobic activity per week, as well as moderate or greater intensity muscle strengthening activities on two or more days a week."  ^d^ Infrequent Insomnia symptoms (yes) defined as difficulty falling or staying asleep most days/every day. | | | | | | | | | | | | | | | | | | | | | |
| ^e^ Restorative sleep (yes) defined as never/some days waking up feeling rested in the past 30 days. | | | | | | | | | | | | | | | | | | | | | |
| ^f^ Depression was defined by the 2019 Field Representative's Manual as a major depressive disorder or as clinical depression that is a common but serious mood disorder. It causes severe symptoms that affect how you feel, think, and handle daily activities, such as sleeping, eating, or working.” | | | | | | | | | | | | | | | | | | | | | |

**Supplemental Figure 2. Prevalence of life satisfaction by race and ethnicity and age, National Health Interview Survey, 2022, (N=25,090)**

Abbreviations: NH (non-Hispanic). Note: Data are presented as column percentages. Percentages may not sum to 100 due to missing rounding. All estimates are weighted for the survey's complex sampling design. All estimates are age-standardized to the U.S. 2020 population, except for age.

Prevalence (%)

|  | Satisfied | Dissatisfied |
| --- | --- | --- |
| Total All | 95.9 | 4.1 |
| Total 18-30 years | 96.3 | 3.7 |
| Total 31-49 years | 96.6 | 3.4 |
| Total ≥ 50 years | 95.3 | 4.7 |
| Hispanic/Latino All | 96.5 | 3.5 |
| Hispanic/Latino 18-30 years | 96.8 | 3.2 |
| Hispanic/Latino 31-49 years | 97.9 | 2.1 |
| Hispanic/Latino ≥ 50 years | 95.8 | 4.2 |
| NH-American Indian/Alaska Native All | 95.3 | 4.7 |
| NH-American Indian/Alaska Native 18-30 years | 92.8 | 7.2 |
| NH-American Indian/Alaska Native 31-49 years | 99.2 | 0.8 |
| NH-American Indian/Alaska Native ≥ 50 years | 94.1 | 5.9 |
| NH-Asian All | 98.1 | 1.9 |
| NH-Asian 18-30 years | 99.3 | 0.7 |
| NH-Asian 31-49 years | 98.3 | 1.7 |
| NH-Asian ≥ 50 years | 97.2 | 2.8 |
| NH-Black/African American All | 95.3 | 4.7 |
| NH-Black/African American 18-30 years | 94.3 | 5.7 |
| NH-Black/African American 31-49 years | 95.9 | 4.1 |
| NH-Black/African American ≥ 50 years | 95.2 | 4.8 |
| NH-Multiracial or other group All | 93.5 | 6.5 |
| NH-Multiracial or other group 18-30 years | 95.7 | 4.3 |
| NH-Multiracial or other group 31-49 years | 92.7 | 7.3 |
| NH-Multiracial or other group ≥ 50 years | 92.5 | 7.5 |
| NH-White All | 95.8 | 4.2 |
| NH-White 18-30 years | 96.3 | 3.7 |
| NH-White 31-49 years | 96.2 | 3.8 |
| NH-White ≥ 50 years | 95.1 | 4.9 |

NH-Other single and multiple races and ‘Other group’ is defined as persons identifying with racial groups not explicitly listed in the standard categories.

Life satisfaction was ascertained based on the questions, 'In general, how satisfied are you with your life? Are you very satisfied, satisfied, dissatisfied, or very dissatisfied?'. Life satisfaction was dichotomized as yes (a response of 'satisfied' or 'very satisfied') vs. no (a response of 'very dissatisfied' or 'dissatisfied').

**Supplemental Figure 3. Prevalence of life satisfaction by race and ethnicity and sex, National Health Interview Survey, 2022, (N=25,090)**

Prevalence (%)

|  | Satisfied | Dissatisfied |
| --- | --- | --- |
| Total All | 95.9 | 4.1 |
| Total Men | 95.8 | 4.2 |
| Total Women | 96.1 | 3.9 |
| Hispanic/Latino All | 96.5 | 3.5 |
| Hispanic/Latino Men | 96.3 | 3.7 |
| Hispanic/Latino Women | 96.8 | 3.2 |
| NH-American Indian/Alaska Native All | 95.3 | 4.7 |
| NH-American Indian/Alaska Native Men | 92.5 | 7.5 |
| NH-American Indian/Alaska Native Women | 97.8 | 2.2 |
| NH-Asian All | 98.1 | 1.9 |
| NH-Asian Men | 98.3 | 1.7 |
| NH-Asian Women | 98 | 2 |
| NH-Black/African American All | 95.3 | 4.7 |
| NH-Black/African American Men | 94.6 | 5.4 |
| NH-Black/African American Women | 95.8 | 4.2 |
| NH-Multiracial or other group All | 93.5 | 6.5 |
| NH-Multiracial or other group Men | 92.2 | 7.8 |
| NH-Multiracial or other group Women | 94.9 | 5.1 |
| NH-White All | 95.8 | 4.2 |
| NH-White Men | 95.8 | 4.2 |
| NH-White Women | 95.8 | 4.2 |

Abbreviations: NH (non-Hispanic)

Note: Data are presented as column percentages or means and standard errors. Percentages may not sum to 100 due to missing rounding. All estimates are weighted for the survey's complex sampling design. All estimates are age-standardized to the U.S. 2020 population, except for age.

NH-Other single and multiple races and ‘Other group’ is defined as persons identifying with racial groups not explicitly listed in the standard categories.

Life satisfaction was ascertained based on the questions, 'In general, how satisfied are you with your life? Are you very satisfied, satisfied, dissatisfied, or very dissatisfied?'. Life satisfaction was dichotomized as yes (a response of 'satisfied' or 'very satisfied') vs. no (a response of 'very dissatisfied' or 'dissatisfied').
